# Using the Excitation/Inhibition Ratio to Optimize the Classification of Autism and Schizophrenia

**DOI:** 10.1101/2022.05.24.22275531

**Authors:** Lavinia Carmen Uscătescu, Christopher J. Hyatt, Jack Dunn, Martin Kronbichler, Vince Calhoun, Silvia Corbera, Kevin Pelphrey, Brian Pittman, Godfrey Pearlson, Michal Assaf

## Abstract

The excitation/inhibition (E/I) ratio has been shown to be imbalanced in individuals diagnosed with autism (AT) or schizophrenia (SZ), relative to neurotypically developed controls (TD). However, the degree of E/I imbalance overlap between SZ and AT has not been extensively compared. Our main objectives were (1) to quantify group differences in the E/I ratio between TD, AT, and SZ, (2) to assess the potential of the E/I ratio for differential diagnosis, and (3) to verify the replicability of our findings in a second, independently-acquired dataset. For each participant, we computed the Hurst exponent (H), an indicator of the E/I ratio, from the timecourses of 53 independent components covering the entire brain. Using Random Forest (RF), we ran a classification analysis using the largerof the two datasets (exploratory dataset; 519 TD, 200 AT, 355 SZ) to determine which of the 53 H would yield the highest performance in classifying SZ and AT. Next, taking the ten most important H from the exploratory dataset and the clinical and phenotypic information collected in the replication dataset (55 TD, 30 AT, 39 SZ), we used RF to compare classification performance using five feature sets: (a) H only; (b) Positive and Negative Syndrome Scale (PANSS) and the Autism Diagnostic Observation Schedule (ADOS) only; (c) PANSS, ADOS, Bermond–Vorst Alexithymia Questionnaire (BVAQ), Empathy Quotient (EQ), and IQ; (d) H, PANSS and ADOS; (e) H, PANSS, ADOS, BVAQ, EQ and IQ. Classification performance using H only was higher in the exploratory dataset (AUC = 84%) compared to the replication dataset (AUC = 72%). In the replication dataset, the highest classification performance was obtained when combining H with PANSS, ADOS, BVAQ, EQ and IQ (i.e., model e; AUC = 83%).

## 1. Introduction

Autism (AT) and schizophrenia (SZ) have been recognized as independent diagnoses since the 1970s (APA, 2013). While AT is primarily characterized by distinct patterns of social communication, and repetitive behaviors, a SZ diagnosis consists of positive (e.g., hallucinations, delusions) and negative (e.g., social withdrawal) symptoms. They also show different onset trajectories (i.e., AT becomes apparent in early childhood, while SZ onset usually occurs in late adolescence or early adulthood), albeit a progression from AT to SZ cannot be excluded (Hsu 2022). However, the heterogeneity of both diagnostic categories (Benkarim et al., 2022; Segal et al., 2023) and their phenotypic overlap (Kästner et al., 2015) can hinder accurate psychiatric diagnosis. More precisely, AT and SZ co-occur in approximately 4% of cases (Lai et al., 2019), and share both social (Oliver et al., 2020) and sensory-motor functioning patterns (Du et al., 2021). Differential diagnosis is additionally hindered by the fact that the common clinical observational and interviews to assess diagnosis-specific symptoms, such as the Autism Diagnostic Observation Schedule (ADOS) and the Positive and Negative Syndrome Scale (PANSS), do not have good specificity (Bastiaansen et al., 2011; Trevisan et al., 2020). The overlap between SZ and AT has led to inquire about the underlying neural mechanisms, and whether these might aid in differential diagnosis (Horien et al., 2022). A recent international machine learning competition aimed to classify AT and typically developed (TD) showed that fMRI data can yield a classification accuracy of ∼ 80% (Traut et al., 2022). Classification accuracies based on structural or functional MRI data can be just as high when distinguishing SZ from TD (de Filippis et al-2022). However, the challenge is greater when tackling diagnosis overlap of heterogeneous nosological categories, such as AT and SZ, that share both genetic variants and neuroimaging patterns (Moreau et al., 2021).

One potential brain-based marker is the excitation/inhibition (E/I) ratio, that has been shown to be different in both AT and SZ compared to TD. This is based on the concerted activity of mostly glutamatergic (i.e., excitatory) and GABAergic (i.e., inhibitory) neurons. The former are the most numerous and project throughout the entire brain, while the latter are fewer and synapse locally (for a comprehensive account of excitatory and inhibitory activity balance in the human brain, see Tatti et al., 2017). A way to estimate the E/I ratio in humans based on non-invasive measures, such as resting state fMRI (rsfMRI), is by computing the Hurst (H) exponent from the acquired timeseries; an increased H values indicates a decreased E/I ratio, and vice-versa (for a review see Campbell & Weber, 2022).

In AT, Rubenstein & Merzenich (2003) first hypothesized that observed sensory processing patterns may result from an increased E/I ratio. Among the evidence they cite is the fact that parietal and cerebellar areas show ∼50% less glutamic acid decarboxylase (GAD), the enzyme that synthesizes the inhibitory neurotransmitter γ - aminobutyric acid (GABA) in AT compared to TD (Fatemi et al., 2002). Additionally, cortical minicolumns, which are functional units composed of GABAergic and glutamatergic neurons processing thalamic inputs, are smaller and more numerous in AT compared to TD (Casanova et al., 2002). A more recent summary specifically points towards the impact of reduced inhibition on cortical and hippocampal functioning in AT (Sohal & Rubenstein, 2019). Whether this E/I imbalance is mainly due to excessive excitatory activity, or deficient inhibitory activity, is not entirely clear (Dickinson, Jones & Milne, 2016; Ford & Crewther, 2016), but recent evidence points to the E/I imbalance in AT being caused by concomitant effect (see Canitano & Palumbi, 2021). Finally, direct evidence for the contribution of an E/I imbalance in AT comes from a study using bumetanide (i.e., a selective NKCC1 chloride importer antagonist, which decreases depolarizing GABA action, to reduce the E/I ratio) in a large cohort of AT children (Juarez-Martinez et al., 2023). These authors reported a decrease in repetitive behaviors following a 91-day bumetanide trial. Another direct link between sensory processing and GABAergic activity in AT has been provided by Huang et al. (2023). These authors used arbaclofen, a GABA type B receptor agonist, to show that auditory repetition suppression was negatively impacted by the drug in TD, but improved in AT.

In SZ, the E/I ratio has also been reported to be imbalanced compared to TD. Post-mortem and genetic evidence (Anticevic & Lisman, 2017), and computational modeling revealed that this imbalance causes hyperconnectivity in association brain areas (Yang et al., 2016). In addition, a review has shown a link between an E/I imbalance and aberrant internal sensory processing in SZ, such as hallucinations (Jardri et al., 2016). Finally, dopamine appears to be crucial in maintaining the E/I balance by modulating the excitability of glutamate and GABAergic neurons, thus contributing crucially to the E/I ration in SZ (Purves-Tyson et al., 2021). Dopaminergic activity, in concerted action with glutamatergic and GABAergic activity, when disrupted, can directly impact memory function and prefrontal connectivity in SZ (please see Winterer & Weinberger, 2004, for an extended account).

Supporting evidence in favor of an E/I imbalance in AT and SZ has been corroborated by animal models and post-mortem human studies, as well as by experimental, genetic, and magnetic resonance spectroscopy studies (MRS) (for a comprehensive review, please see Dickinson et al., 2016). It has been proposed that there are common neuronal pathways underlying E/I imbalance in both AT and SZ (Canitano & Pallagrosi, 2017; Foss-Feig et al., 2017), and that this relies in turn on shared genotype (Gao & Penzes, 2015). However, given the substantial heterogeneity in both AT and SZ (e.g., Segal et al., 2023), it is difficult to ascertain to which extent there is overlap between the two diagnoses.

In recent years, various machine learning approaches have been employed to improve differential diagnosis of mental disorders (Cho et al., 2019). Among these, interpretable models, such as Random Forest (RF), have become increasingly popular due to their transparency, as opposed to the traditional “black box” methods, such as support vector machine (Murdoch et al., 2019; Rudin, 2019). A trade-off between high interpretability and high accuracy is usually considered when opting for a particular classification approach, as highly accurate classifiers usually provide little if any transparency. Among interpretable approaches, RF stands out as an algorithm that can provide both high accuracy and reasonable interpretability (Bhattacharya, 2022). In addition, it can also be used for feature selection based on feature importance in an out-of-sample classification, as we did in the current project (Speiser et al. (2019).

To assess the role of clinical vs. E/I ratio data in classifying AT and SZ, we used two independent datasets and five distinct sets of features (i.e., classification models) comprising phenotypic and clinical assessment scores, the E/I ratio (as indexed by the H exponent) of multiple brain areas, or both. To quantify the E/I ratio based on rsfMRI timeseries, we computed the H of 53 predefined functional brain areas, based on the Neuromark templates (Du et al., 2020). The H exponent has been refined as a reliable computational approximation of synaptic E/I based on extensive physiological and *in silico* studies, (e.g., Trakoshis et al., 2020). For the clinical features we focused on core symptoms assessments: ADOS — measuring AT-related social and communication functioning, and PANSS — measuring SZ-related positive (e.g. delusions, hallucinations) and negative (e.g., social withdrawal) symptoms, and general psychopathology (e.g., attention deficits), an IQ estimate, and two social cognitive measures: EQ — measuring empathy, and BVAQ — measuring alexithymia, both of which have been shown to be different in AT and SZ compared to TD (van’t Wout et al., 2007; Warrier et al., 2018; Kinnaird, Stewart & Tchanturia, 2019). Two rsfMRI datasets were used to test replicability. The first dataset included data from publicly available repositories of either AT or SZ data, and included a relatively large dataset (Du et al., 2022). The second dataset was collected on-site, and included the above-mentioned clinical and phenotypic measures in a smaller dataset. We therefore only tested the out-of-sample replicability of the E/I-based classification model. We believe the use of both datasets holds important advantages. The replication dataset, while consisting of fewer participants, contains both rsfMRI and phenotypic and clinical data. In addition, the AT, SZ and TD in this dataset were collected in the same setting, which precludes the risk of site-related confounds. The larger, exploratory dataset was obtained by sourcing datasets from different online repositories, namely the Autism Brain Imaging Data Exchange (ABIDE I and II) for AT, and the Bipolar-Schizophrenia Network on Intermediate Phenotypes (B-SNIP), the Center for Biomedical Research Excellence (COBRE), the Maryland Psychiatric Research Center (MPRC), and the Function Biomedical Informatics Research Network (FBIRN) for SZ. These datasets had been acquired at various sites with different scanning parameters, and clinical and phenotypical data was also not uniformly available across sites and clinical groups. While this prevented us for doing a full exploration of all the classification models that we were able to test using the smaller dataset, it allowed us to, 1) reduce model complexity and increase model stability (Schultz et al., 2022) in the smaller replication dataset by using only the most important H features (i.e., brain regions) from the larger exploratory dataset, and 2) demonstrate the replicability of the results of this model.

## 2. Methods

### 2.1. Participants

Two independent datasets were used in the current project. An exploratory dataset (Exploratory), based on several publicly-available online datasets (described below), and an internally-collected replication dataset (Replication).

For the Exploratory dataset, we analyzed 519 TD (362 males & 157 females; mean age = 28.49 ± 7.68), 200 AT (180 males & 20 females; mean age = 24.74 ± 6.6), and 355 SZ (245 males & 110 females; mean age = 30.91 ± 7.95) from the previously preprocessed and harmonized dataset used in Du et al. (2022). The participants in Du et al. (2022) had been selected from several data repositories: the AT from ABIDE1 and ABIDE2, and the SZ from BSNIP, COBRE, FBIRN, and MPRC_PK. From the dataset of Du et al. (2022), we chose a subset of participants that closely matched the age (18-35 y.o.) and intelligence quotient (IQ) of the Replication dataset. Note that an estimated IQ > 75 criterion was chosen because it was the inclusion threshold in the BSNIP dataset, where no IQ values were recorded. Because some of the data from the Replication dataset had been previously submitted to data repositories (e.g., ABIDE1) from which the dataset of Du et al. (2022) had been drawn, we ensured that no participants were included in both the Exploratory and Replication datasets, by excluding them from the Exploratory dataset.

For the Replication dataset, participants were recruited via the Olin Neuropsychiatry Research Center (ONRC) and the Yale University School of Medicine and underwent resting state fMRI (rsfMRI) scanning for the current study. We discarded participants with head motion > 10 mm, and those with incomplete phenotypic assessment information, resulting in a final dataset consisting of 55 TD (26 males & 29 females; mean age = 23.86 ± 3.65), 30 AT (25 males & 5 females, mean age = 22.33 ± 3.78), and 39 SZ (31 males & 8 females, mean age = 25.66 ± 3.53). The Replication dataset has been previously used by Hyatt et al. (2020, 2021) and Rabany et al. (2019), and the exclusion criteria we used here were the same: intellectual disability (i.e., estimated IQ < 80), neurological disorder (e.g., epilepsy), current drug use as indicated by pre-scanning interview and urine test, incompatibility with MRI safety measures (e.g., metal implants), and a history of psychiatric diagnoses in TD.

### 2.2. Clinical and phenotypical assessment

Clinical and phenotypic data was collected from the Replication dataset, but was not consistently available for the Exploratory dataset. The clinical assessment focused on assessing psychotic and autistic features. The severity of psychotic symptoms was assessed using the Positive and Negative Syndrome Scale (PANSS; Kay et al., 1987) in both the AT and SZ group. The PANSS scores can be interpreted along three subscales: positive symptoms, reflecting the severity of hallucinations and delusions; negative symptoms, reflecting the severity of blunted affect and anhedonia, and a general subscale quantifying other psychopathology such as poor attention and lack of insight. The ADOS, module 4 (Lord et al., 2000) was administered to all participants and the total score was used in this study to confirm or rule out an autism diagnosis and quantify autistic social communication characteristics. The Structured clinical interview for DSM-IV-TR Axis I disorders (SCID; First & Gibbon, 2004) to confirm a SZ diagnosis and the absence of any Axis I diagnoses in TD. Estimated IQ was calculated for the entire dataset using the Vocabulary and Block Design subtests of the *Wechsler Scale of Adult Intelligence-III* (WAIS-III; Wechsler, 1997; Sattler and Ryan, 1999). Additionally, all participants completed the Empathizing Quotient (EQ; Wakabayashi, Baron-Cohen & Wheelwright, 2006) which measures general empathy including both the affective and cognitive empathy components; the Bermond–Vorst Alexithymia Questionnaire (BVAQ; Vorst & Bermond, 2001), whose subscores are computed along five distinct dimensions: “verbalizing” reflects one’s propensity to talk about one’s feelings; “identifying” reflects the extent to which one is able to accurately define one’s emotional states; “analyzing” quantifies the extent to which one seeks to understand the reason for one’s emotions; “fantasizing” quantifies one’s tendency to day-dream, and “emotionalizing” reflects the extent to which a person is emotionally aroused by emotion-inducing events. Descriptive statistics and group comparisons of the clinical and phenotypic data are given in Table 1.

**Table 1.**
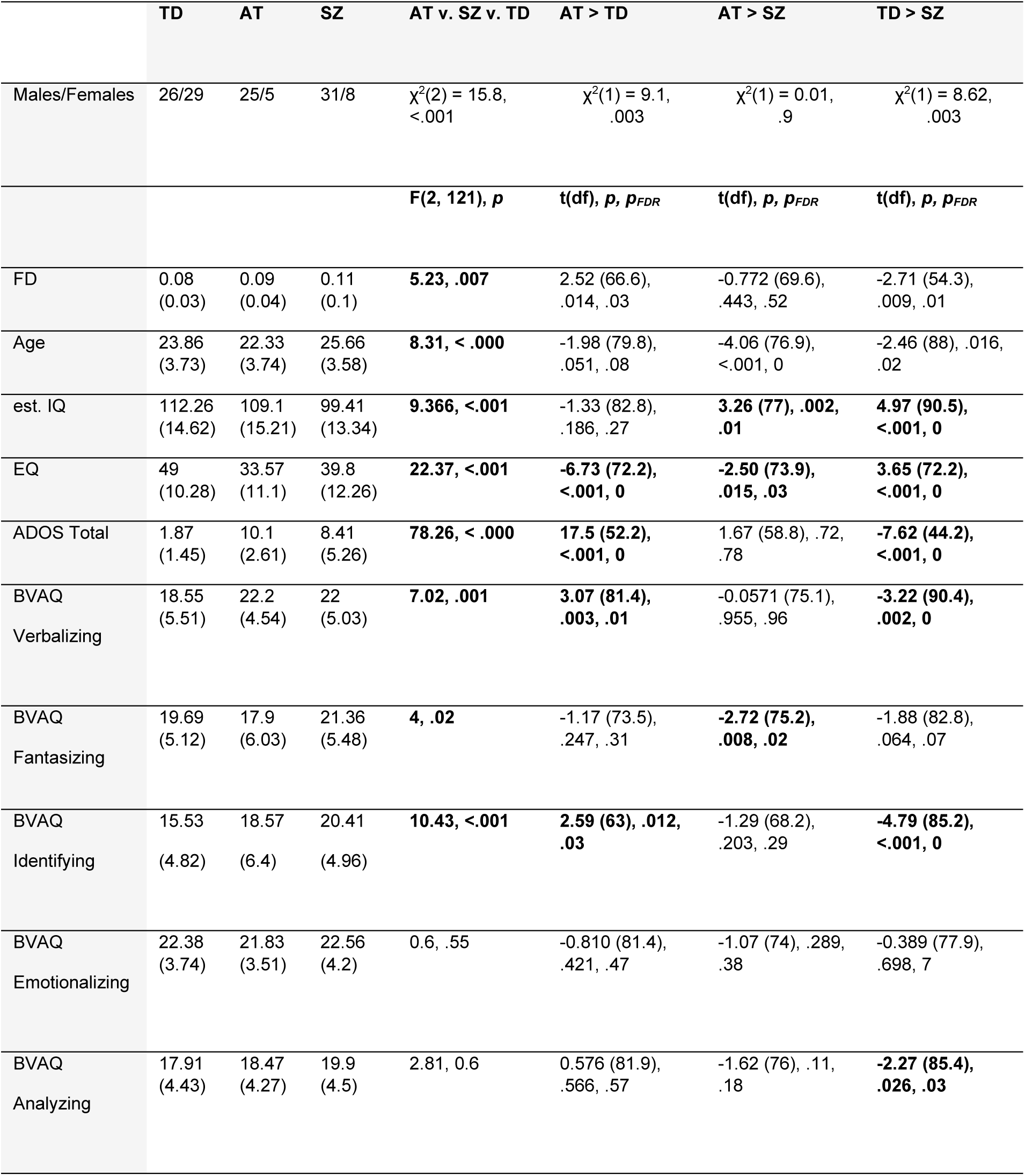
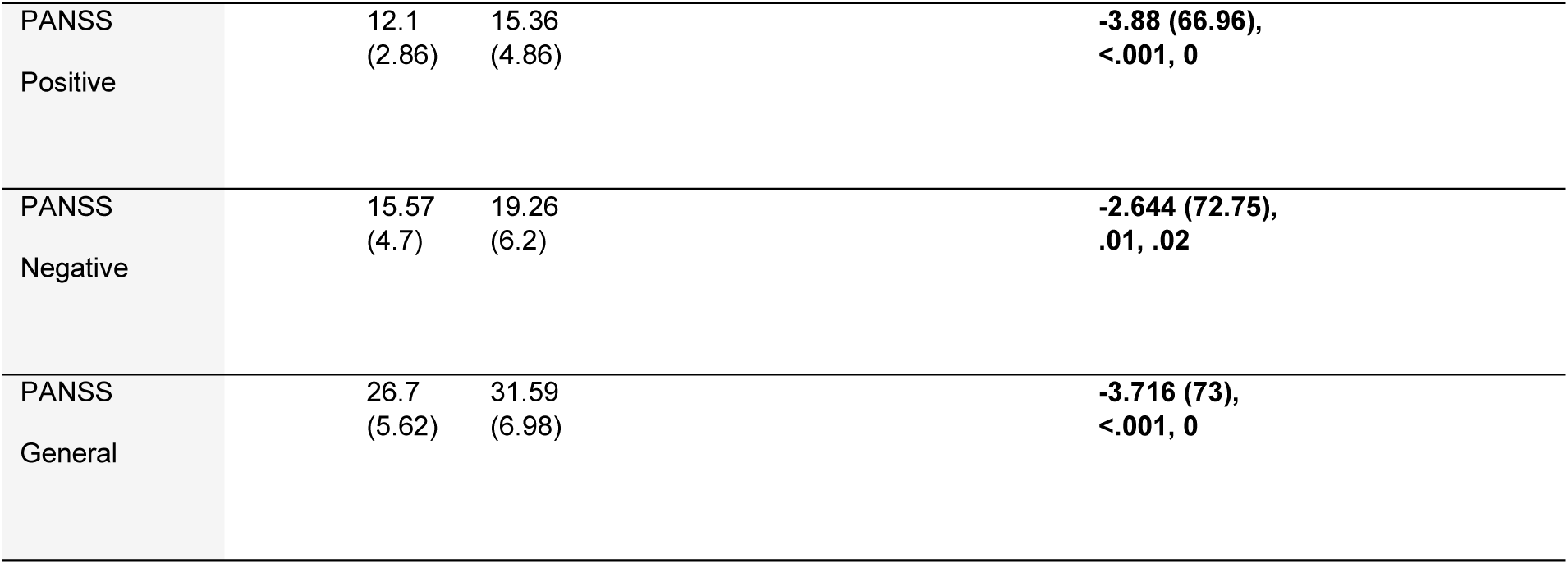
Means and standard deviations (in parentheses) of demographics, phenotypic and clinical scores for all three groups of the Replication dataset: est. IQ = estimated Intelligence Quotient; EQ = Empathy Quotient; ADOS = Autism Diagnostic Observation Schedule module 4; BVAQ = Bermond– Vorst Alexithymia Questionnaire; FD = framewise displacement. Group statistics are shown in the last four columns. Pairwise comparisons were performed using Welch two-samples t test. Both uncorrected (i.e., *p*) and false discovery rate corrected (i.e., *p*FDR) *p* values are shown.

### 2.3. Imaging data acquisition and preprocessing

The preprocessing steps of the rsfMRI data, using the SPM toolbox, of the Exploratory dataset were extensively described in Du et al. (2022). In short, the first few volumes were discarded, then rigid-body motion correction and slice-timing correction were performed. Finally, the data were normalized, resampled to 3 mm^3^ isotropic voxels, and smoothed with a 6 mm FWHM Gaussian kernel. Prior to preprocessing, the effects of age, gender, site acquisition, and interactions between age and site, and gender and site were regressed from the gray matter volumes of each voxel, to ensure between-site harmonization; this procedure was detailed in Du et al. (2022).

For the Replication dataset, rsfMRI scans lasted 7.5 min and were collected using a Siemens Skyra 3 T scanner at the ONRC. Participants lay still, with eyes open, while fixating a centrally presented cross. Blood oxygenation level dependent (BOLD) signal was obtained with a T2*-weighted echo planar imaging (EPI) sequence: TR = 475 ms, TE = 30 ms, flip angle = 60 deg, 48 slices, multiband (8), interleaved slice order, 3 mm^3^ voxels. Neuroimaging data were preprocessed using SPM8 (www.fil.ion.ucl.ac.uk/spm/software/spm8/). Each dataset was realigned to the first T2* image using the INRIAlign toolbox (https://www-sop.inria.fr/epidaure/Collaborations/IRMf/INRIAlign.html), coregistered to their corresponding high signal-to-noise single-band reference image (sbREF; Glasser et al., 2013), spatially normalized to the Montreal Neurological Institute (MNI) standard template (Friston et al., 1995), and spatially smoothed (6 mm^3^). Finally, framewise displacement (FD) motion parameters were computed according to the FSL library algorithm (Jenkinson et al., 2012), and the mean FD value for each run was used as a covariate in group analyses.

### 2.4. Data analysis

For both the Exploratory and Replication dataset, we ran a fully automated independent component analysis (ICA) on the preprocessed fMRI data using the Group ICA for fMRI Toolbox (GIFT v4.0c; https://trendscenter.org/software/gift/; Calhoun et al., 2001) to define functional brain regions. The 53 replicable independent component (IC) templates from the NeuroMark pipeline (Du et al., 2020) were used to estimate participant-specific, spatially-independent components using a spatially-constrained ICA algorithm (Du et al., 2018). A complete list of the NeuroMark IC templates, arranged into seven functional domains, and peak MNI coordinates for each IC template are given in Table 2 and illustrated in Supplement Figure 1. After detrending and despiking using 3dDespike (AFNI, 1995), we extracted one Hurst exponent (H), an estimate of the E/I ratio, from each of the resulting 53 IC timecourses of each participant.

**Table 2.**
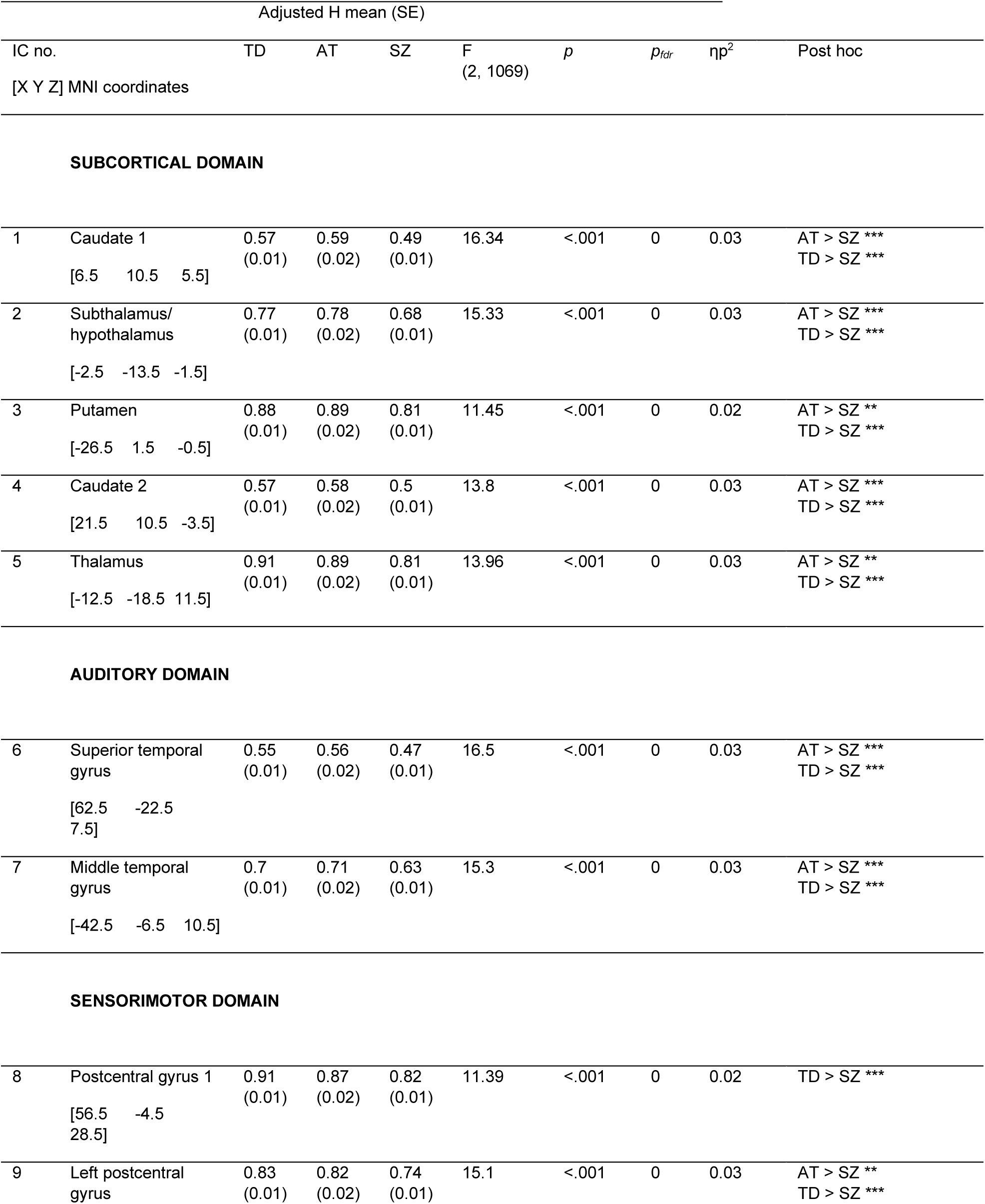
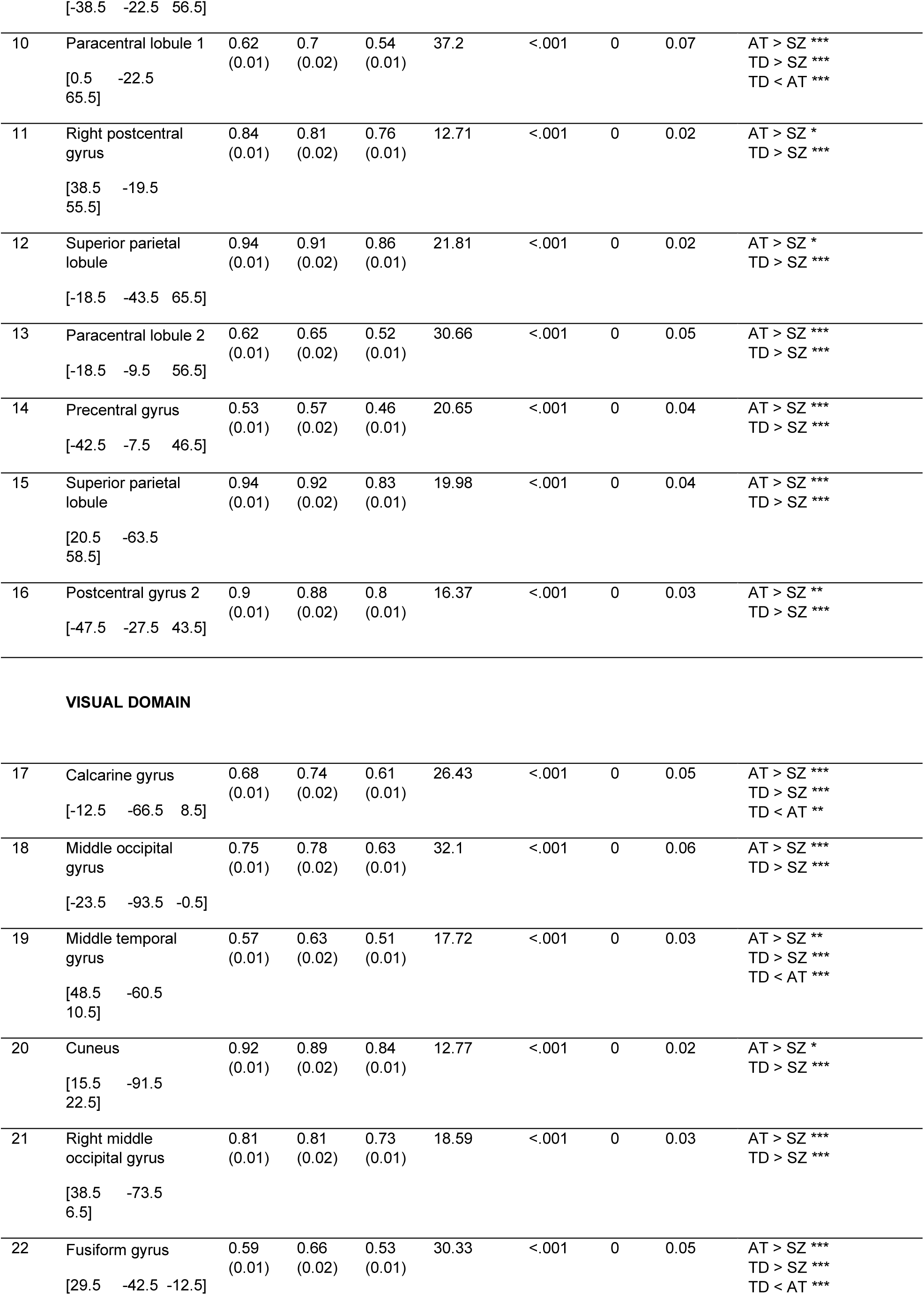
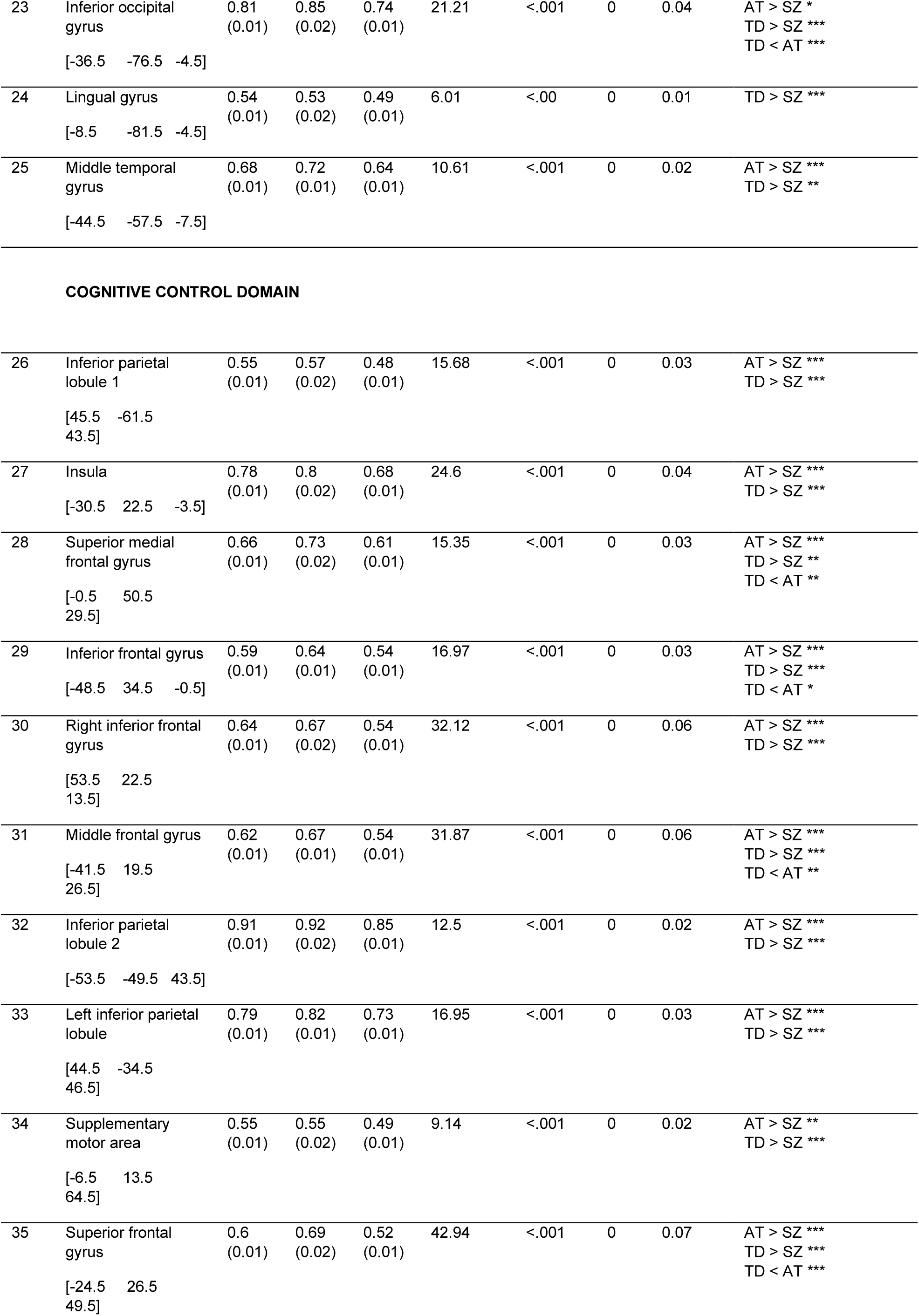
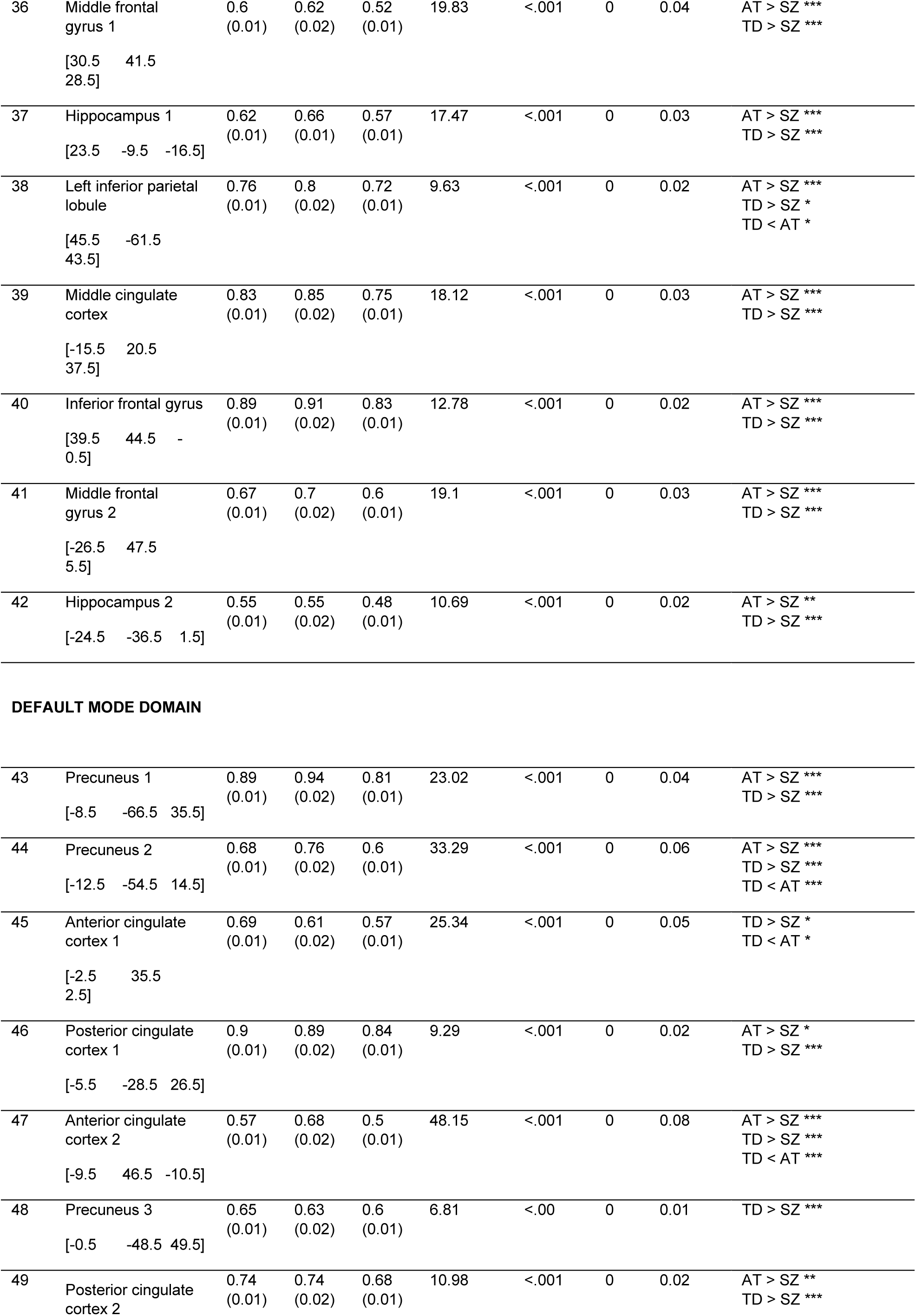
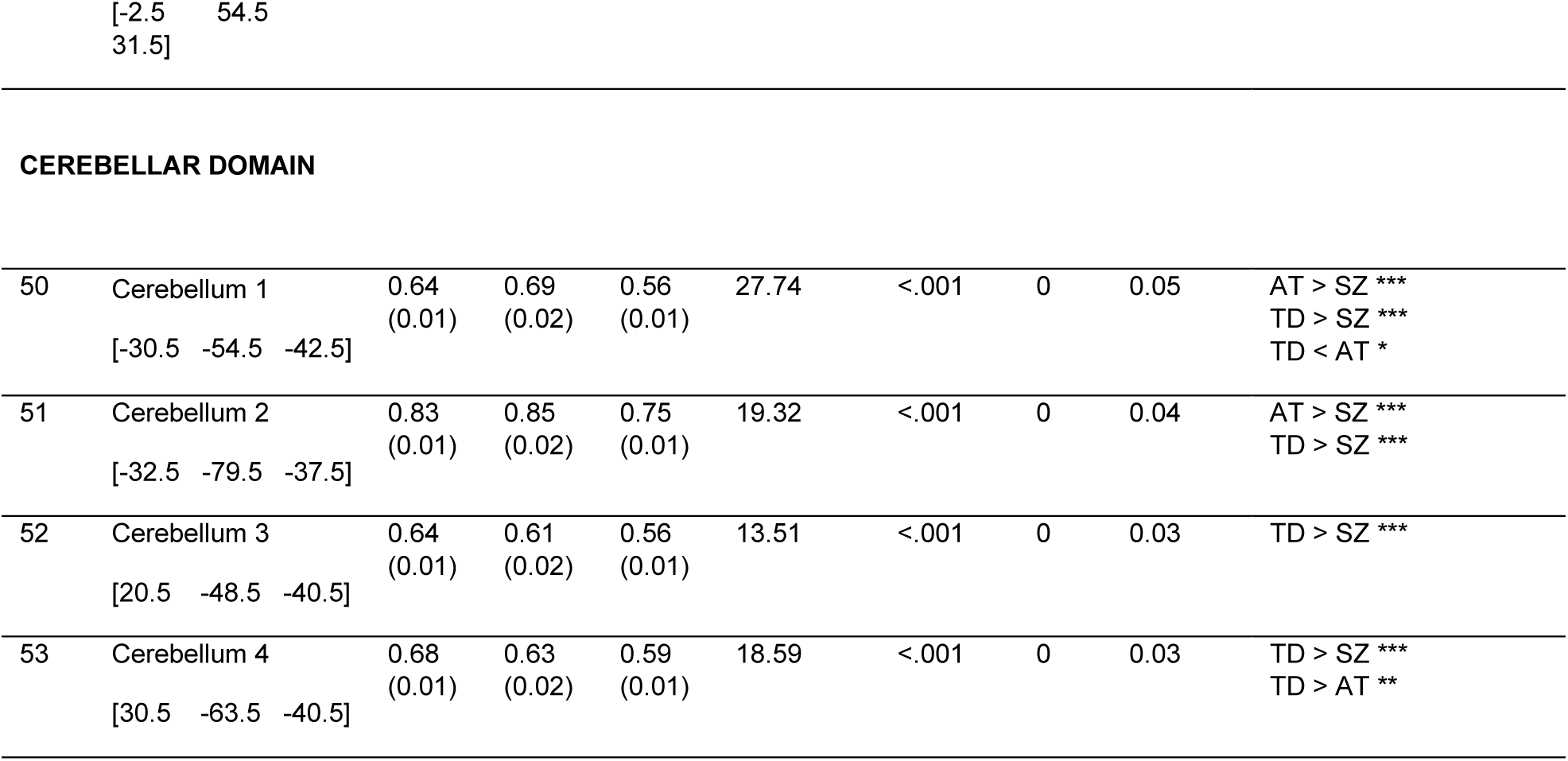
Group differences in Hurst exponent (H) per component, in the Exploratory dataset, computed using ANCOVA with age and sex as covariates. Multiple comparison correction of the ANCOVA p values was performed using false discovery rate (fdr). The ANCOVA effect size was calculated using partial η^2^. Post hoc tests were ran using Tukey’s HSD Test for multiple comparisons with C.I. = 95%. * *p* < .05; ** *p* < .01; *** *p* < .001

|  |  | Adjusted H mean (SE) |  |  |  |  |  |  |  |
| --- | --- | --- | --- | --- | --- | --- | --- | --- | --- |
| IC no. |  | TD | AT | SZ | F<br>(2, 1069) | *p* | *p*<sub>fdr</sub> | η*p*<sup>2</sup> | Post hoc |
| [X Y Z] MNI coordinates |  |  |  |  |  |  |  |  |  |
| SUBCORTICAL DOMAIN |  |  |  |  |  |  |  |  |  |
| 1 | Caudate 1<br>[6.5 10.5 5.5] | 0.57<br>(0.01) | 0.59<br>(0.02) | 0.49<br>(0.01) | 16.34 | <.001 | 0 | 0.03 | AT > SZ ***<br>TD > SZ *** |
| 2 | Subthalamus/<br>hypothalamus<br>[-2.5 -13.5 -1.5] | 0.77<br>(0.01) | 0.78<br>(0.02) | 0.68<br>(0.01) | 15.33 | <.001 | 0 | 0.03 | AT > SZ ***<br>TD > SZ *** |
| 3 | Putamen<br>[-26.5 1.5 -0.5] | 0.88<br>(0.01) | 0.89<br>(0.02) | 0.81<br>(0.01) | 11.45 | <.001 | 0 | 0.02 | AT > SZ **<br>TD > SZ *** |
| 4 | Caudate 2<br>[21.5 10.5 -3.5] | 0.57<br>(0.01) | 0.58<br>(0.02) | 0.5<br>(0.01) | 13.8 | <.001 | 0 | 0.03 | AT > SZ ***<br>TD > SZ *** |
| 5 | Thalamus<br>[-12.5 -18.5 11.5] | 0.91<br>(0.01) | 0.89<br>(0.02) | 0.81<br>(0.01) | 13.96 | <.001 | 0 | 0.03 | AT > SZ **<br>TD > SZ *** |
| AUDITORY DOMAIN |  |  |  |  |  |  |  |  |  |
| 6 | Superior temporal<br>gyrus<br>[62.5 -22.5<br>7.5] | 0.55<br>(0.01) | 0.56<br>(0.02) | 0.47<br>(0.01) | 16.5 | <.001 | 0 | 0.03 | AT > SZ ***<br>TD > SZ *** |
| 7 | Middle temporal<br>gyrus<br>[-42.5 -6.5 10.5] | 0.7<br>(0.01) | 0.71<br>(0.02) | 0.63<br>(0.01) | 15.3 | <.001 | 0 | 0.03 | AT > SZ ***<br>TD > SZ *** |
| SENSORIMOTOR DOMAIN |  |  |  |  |  |  |  |  |  |
| 8 | Postcentral gyrus 1<br>[56.5 -4.5<br>28.5] | 0.91<br>(0.01) | 0.87<br>(0.02) | 0.82<br>(0.01) | 11.39 | <.001 | 0 | 0.02 | TD > SZ *** |
| 9 | Left postcentral<br>gyrus | 0.83<br>(0.01) | 0.82<br>(0.02) | 0.74<br>(0.01) | 15.1 | <.001 | 0 | 0.03 | AT > SZ **<br>TD > SZ *** |
| 10 | Paracentral lobule 1<br>[0.5 -22.5 65.5] | 0.62<br>(0.01) | 0.7<br>(0.02) | 0.54<br>(0.01) | 37.2 | <.001 | 0 | 0.07 | AT > SZ ***<br>TD > SZ ***<br>TD < AT *** |
| 11 | Right postcentral gyrus<br>[38.5 -19.5 55.5] | 0.84<br>(0.01) | 0.81<br>(0.02) | 0.76<br>(0.01) | 12.71 | <.001 | 0 | 0.02 | AT > SZ *<br>TD > SZ *** |
| 12 | Superior parietal lobule<br>[-18.5 -43.5 65.5] | 0.94<br>(0.01) | 0.91<br>(0.02) | 0.86<br>(0.01) | 21.81 | <.001 | 0 | 0.02 | AT > SZ *<br>TD > SZ *** |
| 13 | Paracentral lobule 2<br>[-18.5 -9.5 56.5] | 0.62<br>(0.01) | 0.65<br>(0.02) | 0.52<br>(0.01) | 30.66 | <.001 | 0 | 0.05 | AT > SZ ***<br>TD > SZ *** |
| 14 | Precentral gyrus<br>[-42.5 -7.5 46.5] | 0.53<br>(0.01) | 0.57<br>(0.02) | 0.46<br>(0.01) | 20.65 | <.001 | 0 | 0.04 | AT > SZ ***<br>TD > SZ *** |
| 15 | Superior parietal lobule<br>[20.5 -63.5 58.5] | 0.94<br>(0.01) | 0.92<br>(0.02) | 0.83<br>(0.01) | 19.98 | <.001 | 0 | 0.04 | AT > SZ ***<br>TD > SZ *** |
| 16 | Postcentral gyrus 2<br>[-47.5 -27.5 43.5] | 0.9<br>(0.01) | 0.88<br>(0.02) | 0.8<br>(0.01) | 16.37 | <.001 | 0 | 0.03 | AT > SZ **<br>TD > SZ *** |
| VISUAL DOMAIN |  |  |  |  |  |  |  |  |  |
| 17 | Calcarine gyrus<br>[-12.5 -66.5 8.5] | 0.68<br>(0.01) | 0.74<br>(0.02) | 0.61<br>(0.01) | 26.43 | <.001 | 0 | 0.05 | AT > SZ ***<br>TD > SZ ***<br>TD < AT ** |
| 18 | Middle occipital gyrus<br>[-23.5 -93.5 -0.5] | 0.75<br>(0.01) | 0.78<br>(0.02) | 0.63<br>(0.01) | 32.1 | <.001 | 0 | 0.06 | AT > SZ ***<br>TD > SZ *** |
| 19 | Middle temporal gyrus<br>[48.5 -60.5 10.5] | 0.57<br>(0.01) | 0.63<br>(0.02) | 0.51<br>(0.01) | 17.72 | <.001 | 0 | 0.03 | AT > SZ **<br>TD > SZ ***<br>TD < AT *** |
| 20 | Cuneus<br>[15.5 -91.5 22.5] | 0.92<br>(0.01) | 0.89<br>(0.02) | 0.84<br>(0.01) | 12.77 | <.001 | 0 | 0.02 | AT > SZ *<br>TD > SZ *** |
| 21 | Right middle occipital gyrus<br>[38.5 -73.5 6.5] | 0.81<br>(0.01) | 0.81<br>(0.02) | 0.73<br>(0.01) | 18.59 | <.001 | 0 | 0.03 | AT > SZ ***<br>TD > SZ *** |
| 22 | Fusiform gyrus<br>[29.5 -42.5 -12.5] | 0.59<br>(0.01) | 0.66<br>(0.02) | 0.53<br>(0.01) | 30.33 | <.001 | 0 | 0.05 | AT > SZ ***<br>TD > SZ ***<br>TD < AT *** |
| 23 | Inferior occipital gyrus<br>[-36.5 -76.5 -4.5] | 0.81<br>(0.01) | 0.85<br>(0.02) | 0.74<br>(0.01) | 21.21 | <.001 | 0 | 0.04 | AT > SZ *<br>TD > SZ ***<br>TD < AT *** |
| 24 | Lingual gyrus<br>[-8.5 -81.5 -4.5] | 0.54<br>(0.01) | 0.53<br>(0.02) | 0.49<br>(0.01) | 6.01 | <.00 | 0 | 0.01 | TD > SZ *** |
| 25 | Middle temporal gyrus<br>[-44.5 -57.5 -7.5] | 0.68<br>(0.01) | 0.72<br>(0.01) | 0.64<br>(0.01) | 10.61 | <.001 | 0 | 0.02 | AT > SZ ***<br>TD > SZ ** |
| COGNITIVE CONTROL DOMAIN |  |  |  |  |  |  |  |  |  |
| 26 | Inferior parietal lobule 1<br>[45.5 -61.5 43.5] | 0.55<br>(0.01) | 0.57<br>(0.02) | 0.48<br>(0.01) | 15.68 | <.001 | 0 | 0.03 | AT > SZ ***<br>TD > SZ *** |
| 27 | Insula<br>[-30.5 22.5 -3.5] | 0.78<br>(0.01) | 0.8<br>(0.02) | 0.68<br>(0.01) | 24.6 | <.001 | 0 | 0.04 | AT > SZ ***<br>TD > SZ *** |
| 28 | Superior medial frontal gyrus<br>[-0.5 50.5 29.5] | 0.66<br>(0.01) | 0.73<br>(0.02) | 0.61<br>(0.01) | 15.35 | <.001 | 0 | 0.03 | AT > SZ ***<br>TD > SZ **<br>TD < AT ** |
| 29 | Inferior frontal gyrus<br>[-48.5 34.5 -0.5] | 0.59<br>(0.01) | 0.64<br>(0.01) | 0.54<br>(0.01) | 16.97 | <.001 | 0 | 0.03 | AT > SZ ***<br>TD > SZ ***<br>TD < AT * |
| 30 | Right inferior frontal gyrus<br>[53.5 22.5 13.5] | 0.64<br>(0.01) | 0.67<br>(0.02) | 0.54<br>(0.01) | 32.12 | <.001 | 0 | 0.06 | AT > SZ ***<br>TD > SZ *** |
| 31 | Middle frontal gyrus<br>[-41.5 19.5 26.5] | 0.62<br>(0.01) | 0.67<br>(0.01) | 0.54<br>(0.01) | 31.87 | <.001 | 0 | 0.06 | AT > SZ ***<br>TD > SZ ***<br>TD < AT ** |
| 32 | Inferior parietal lobule 2<br>[-53.5 -49.5 43.5] | 0.91<br>(0.01) | 0.92<br>(0.02) | 0.85<br>(0.01) | 12.5 | <.001 | 0 | 0.02 | AT > SZ ***<br>TD > SZ *** |
| 33 | Left inferior parietal lobule<br>[44.5 -34.5 46.5] | 0.79<br>(0.01) | 0.82<br>(0.01) | 0.73<br>(0.01) | 16.95 | <.001 | 0 | 0.03 | AT > SZ ***<br>TD > SZ *** |
| 34 | Supplementary motor area<br>[-6.5 13.5 64.5] | 0.55<br>(0.01) | 0.55<br>(0.02) | 0.49<br>(0.01) | 9.14 | <.001 | 0 | 0.02 | AT > SZ **<br>TD > SZ *** |
| 35 | Superior frontal gyrus<br>[-24.5 26.5 49.5] | 0.6<br>(0.01) | 0.69<br>(0.02) | 0.52<br>(0.01) | 42.94 | <.001 | 0 | 0.07 | AT > SZ ***<br>TD > SZ ***<br>TD < AT *** |
| 36 | Middle frontal gyrus 1<br>[30.5 41.5<br>28.5] | 0.6<br>(0.01) | 0.62<br>(0.02) | 0.52<br>(0.01) | 19.83 | <.001 | 0 | 0.04 | AT > SZ ***<br>TD > SZ *** |
| 37 | Hippocampus 1<br>[23.5 -9.5 -16.5] | 0.62<br>(0.01) | 0.66<br>(0.01) | 0.57<br>(0.01) | 17.47 | <.001 | 0 | 0.03 | AT > SZ ***<br>TD > SZ *** |
| 38 | Left inferior parietal lobule<br>[45.5 -61.5<br>43.5] | 0.76<br>(0.01) | 0.8<br>(0.02) | 0.72<br>(0.01) | 9.63 | <.001 | 0 | 0.02 | AT > SZ ***<br>TD > SZ *<br>TD < AT * |
| 39 | Middle cingulate cortex<br>[-15.5 20.5<br>37.5] | 0.83<br>(0.01) | 0.85<br>(0.02) | 0.75<br>(0.01) | 18.12 | <.001 | 0 | 0.03 | AT > SZ ***<br>TD > SZ *** |
| 40 | Inferior frontal gyrus<br>[39.5 44.5 -<br>0.5] | 0.89<br>(0.01) | 0.91<br>(0.02) | 0.83<br>(0.01) | 12.78 | <.001 | 0 | 0.02 | AT > SZ ***<br>TD > SZ *** |
| 41 | Middle frontal gyrus 2<br>[-26.5 47.5<br>5.5] | 0.67<br>(0.01) | 0.7<br>(0.02) | 0.6<br>(0.01) | 19.1 | <.001 | 0 | 0.03 | AT > SZ ***<br>TD > SZ *** |
| 42 | Hippocampus 2<br>[-24.5 -36.5 1.5] | 0.55<br>(0.01) | 0.55<br>(0.02) | 0.48<br>(0.01) | 10.69 | <.001 | 0 | 0.02 | AT > SZ **<br>TD > SZ *** |
| DEFAULT MODE DOMAIN |  |  |  |  |  |  |  |  |  |
| 43 | Precuneus 1<br>[-8.5 -66.5 35.5] | 0.89<br>(0.01) | 0.94<br>(0.02) | 0.81<br>(0.01) | 23.02 | <.001 | 0 | 0.04 | AT > SZ ***<br>TD > SZ *** |
| 44 | Precuneus 2<br>[-12.5 -54.5 14.5] | 0.68<br>(0.01) | 0.76<br>(0.02) | 0.6<br>(0.01) | 33.29 | <.001 | 0 | 0.06 | AT > SZ ***<br>TD > SZ ***<br>TD < AT *** |
| 45 | Anterior cingulate cortex 1<br>[-2.5 35.5<br>2.5] | 0.69<br>(0.01) | 0.61<br>(0.02) | 0.57<br>(0.01) | 25.34 | <.001 | 0 | 0.05 | TD > SZ *<br>TD < AT * |
| 46 | Posterior cingulate cortex 1<br>[-5.5 -28.5 26.5] | 0.9<br>(0.01) | 0.89<br>(0.02) | 0.84<br>(0.01) | 9.29 | <.001 | 0 | 0.02 | AT > SZ *<br>TD > SZ *** |
| 47 | Anterior cingulate cortex 2<br>[-9.5 46.5 -10.5] | 0.57<br>(0.01) | 0.68<br>(0.02) | 0.5<br>(0.01) | 48.15 | <.001 | 0 | 0.08 | AT > SZ ***<br>TD > SZ ***<br>TD < AT *** |
| 48 | Precuneus 3<br>[-0.5 -48.5 49.5] | 0.65<br>(0.01) | 0.63<br>(0.02) | 0.6<br>(0.01) | 6.81 | <.00 | 0 | 0.01 | TD > SZ *** |
| 49 | Posterior cingulate cortex 2 | 0.74<br>(0.01) | 0.74<br>(0.02) | 0.68<br>(0.01) | 10.98 | <.001 | 0 | 0.02 | AT > SZ **<br>TD > SZ *** |
| CEREBELLAR DOMAIN |  |  |  |  |  |  |  |  |  |
| 50 | Cerebellum 1<br>[-30.5 -54.5 -42.5] | 0.64<br>(0.01) | 0.69<br>(0.02) | 0.56<br>(0.01) | 27.74 | <.001 | 0 | 0.05 | AT > SZ ***<br>TD > SZ ***<br>TD < AT * |
| 51 | Cerebellum 2<br>[-32.5 -79.5 -37.5] | 0.83<br>(0.01) | 0.85<br>(0.02) | 0.75<br>(0.01) | 19.32 | <.001 | 0 | 0.04 | AT > SZ ***<br>TD > SZ *** |
| 52 | Cerebellum 3<br>[20.5 -48.5 -40.5] | 0.64<br>(0.01) | 0.61<br>(0.02) | 0.56<br>(0.01) | 13.51 | <.001 | 0 | 0.03 | TD > SZ *** |
| 53 | Cerebellum 4<br>[30.5 -63.5 -40.5] | 0.68<br>(0.01) | 0.63<br>(0.02) | 0.59<br>(0.01) | 18.59 | <.001 | 0 | 0.03 | TD > SZ ***<br>TD > AT ** |

We estimated H using the *nonfractal* MATLAB toolbox (https://github.com/wonsang/nonfractal; You et al., 2012). Specifically, we used the function *bfn_mfin_ml.m* with the “filter” argument set to “haar” and the “ub” (upper bound) and “lb” (lower bound) arguments set to [1.5,10] and [-0.5,0], respectively, as previously recommended by Trakoshis et al. (2020).

All the other statistical analyses were performed with R 4.1.1. These included a one-way analysis of covariance (ANCOVA), Tuckey post-hoc, and two-sided two-sample Welch t tests.

#### Classification/Discriminant Analyses

A crucial aspect of classification algorithms in neuroimaging is sample size. It has been shown that larger sample sizes lead to more accurate estimates when brain-behavior relationships are investigated via traditional statistical approaches (Marek et al., 2022). While this seems to be generally true also for machine learning (Schultz et al., 2022), the relationship between sample size and classification accuracy does not appear to be entirely linear (e.g., Flint et al., 2021). For this reason, in the current project, we established the initial classification accuracy of our brain-based classification model in the largest of our two datasets.

First, using the Exploratory dataset, we classified the AT and SZ participants using a random forest (RF) algorithm, as implemented in the Interpretable Artificial Intelligence (IAI) toolbox (https://www.interpretable.ai/) and accessed through R 4.2.1 (R Core Team, 2018). The feature set consisted of the 53 H exponents of each participant. We ran 100 randomized sample splits and averaged the model performance metrics that we obtained for each of the splits to obtain a final classification performance index (i.e., area under the curve/AUC, sensitivity and specificity). From each group, with each new split, 50% of the data was randomly allocated to the test, and the rest to the train group. We used the AT sample as reference for calculating sensitivity and specificity. Following the classification, we selected the 10 ICs with the highest feature importance of H (figure/table x) as a simplified feature set for use in RF classification of the Replication dataset in the next step. Next, using the Replication dataset, we classified the AT and SZ participants using the same algorithm and toolbox, with 100 splits and 50% randomized allocation of data into the train and test groups. Five models were used for the RF classification in this case, containing the following features: (a) E/I model: the H only values of the 10 ICs ranked as most important by the RF classification in the Exploratory dataset; (b) symptoms only model: PANSS 3 factor scores and ADOS total scores; (c) symptoms and cognitive model: PANSS, ADOS, EQ, BVAQ, and IQ scores; (d) E/I and symptoms model: the 10 H from model (a) plus the PANSS and ADOS scores, and (e) E/I, symptoms and cognitive model: the 10 H from model (a) plus the PANSS, ADOS, EQ, BVAQ, and IQ scores. Similar to the previous step, we used the AT sample as reference to calculate sensitivity and specificity, and each final model performance index (i.e., AUC, sensitivity and specificity) was obtained by averaging the respective model performance metric over the 100 sample splits. Misclassification for each participant for each model was calculated as the ratio of number of times each participant was misclassified, to the total times s/he was allocated to a test set.

## 3. Results

### 3.1. Group differences in demographic, clinical and phenotypic assessment

In the Exploratory dataset, there were significant group differences differences on age (F (2) = 42.45, *p* < .001), and sex distribution (χ^2^ (2) = 35.156, *p* <.001).

Data for the Replication dataset, including statistical tests, are presented in Table 1. There were significant differences in estimated IQ, age, sex and FD, and therefore these parameters were used as covariates in further group analyses. Regarding core symptom assessments, the AT and SZ groups did not significantly differ in their social and communication skills, as indicated by the ADOS scores, but the PANSS scores on all three domains (i.e., positive and negative symptoms, and general psychopathology) were significantly elevated in SZ compared to AT. For social tasks, BVAQ-Fantasizing was significantly decreased in AT compared to SZ. Empathy was significantly decreased in both AT and SZ compared to TD, and in AT compared to SZ.

### 3.2. Group differences in H in the Exploratory dataset

The ANOVA results testing group differences in H values are given in Table 2. While all areas showed significant group differences, the largest effect size (i.e., η^2^ >= 0.06) was found for: the paracentral lobule (i.e., ICs no. 10 and 13; belonging to the Sensorimotor domain), the calcarine gyrus, middle occipital gyrus, fusiform gyrus, and inferior occipital gyrus (i.e., ICs no. 17, 18, 20, 23; in the Visual network), the insula, and the superior, middle and right inferior frontal gyrus (i.e., ICs no. 27, 30, 31, 35; in Cognitive control domain), the precuneus and anterior cingulate cortex (i.e., IC no. 43, 44, 47; Default Mode network), and one area of the Cerebellar domain (i.e., IC no. 50).

### 3.3. Group differences in H in the Replication dataset

The ANCOVA results showing group differences in H values from each component are given in Table 3. The areas most sensitive to overall group differences, after controlling for age, sex, IQ, and FD, were the left and right postcentral gyrus and paracentral lobule (i.e., IC no. 9, 11, and 10 respectively, part of the Sensorimotor network), and the calcarine gyrus, middle occipital gyrus, middle temporal gyrus, inferior occipital gyrus, and lingual gyrus (i.e., IC no. 17, 18, 19, 23 and 24 part of the Visual network). The supplementary motor area (i.e., IC no. 34), associated with the Cognitive Control domain, also yielded significant group differences.

**Table 3.**
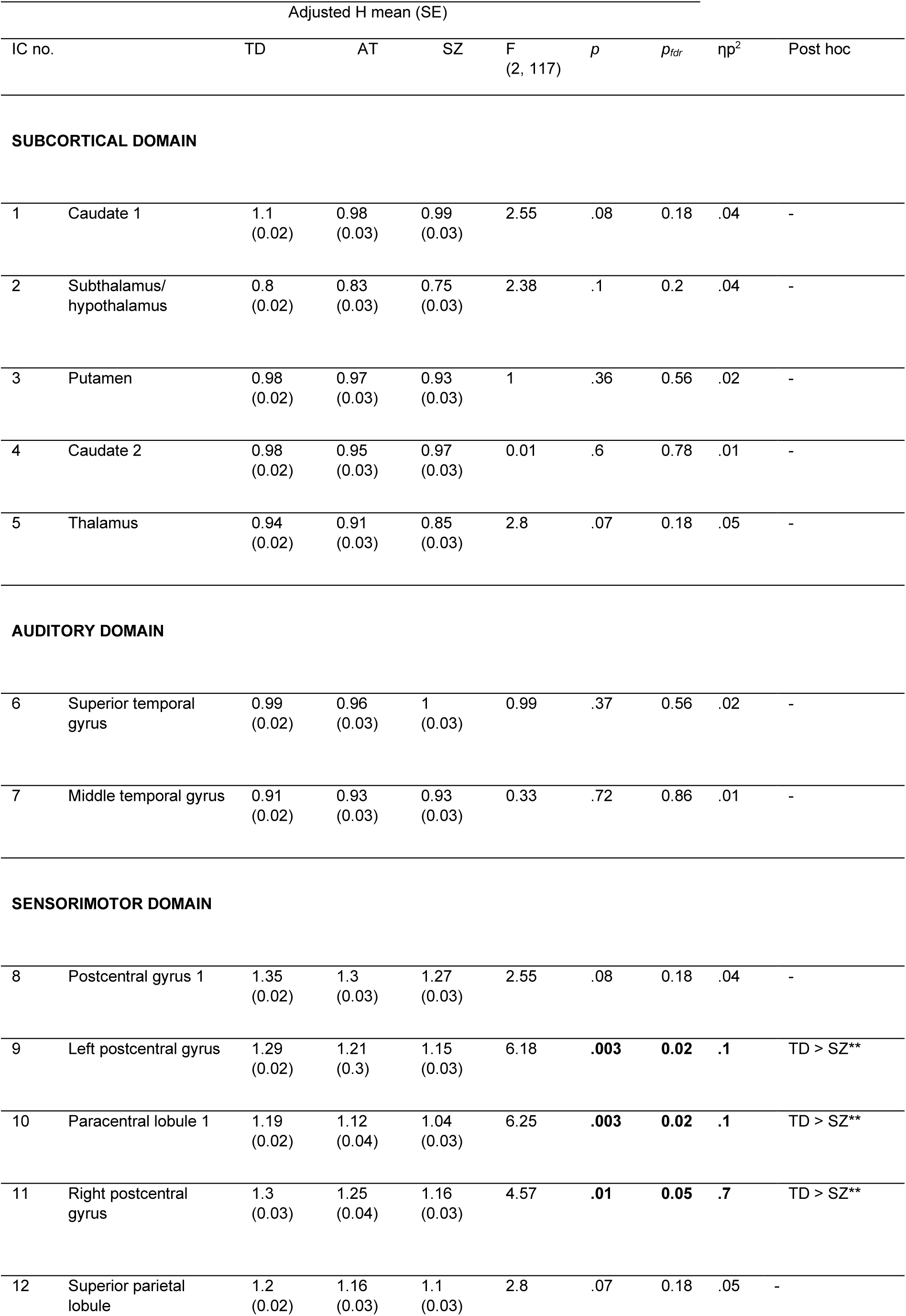
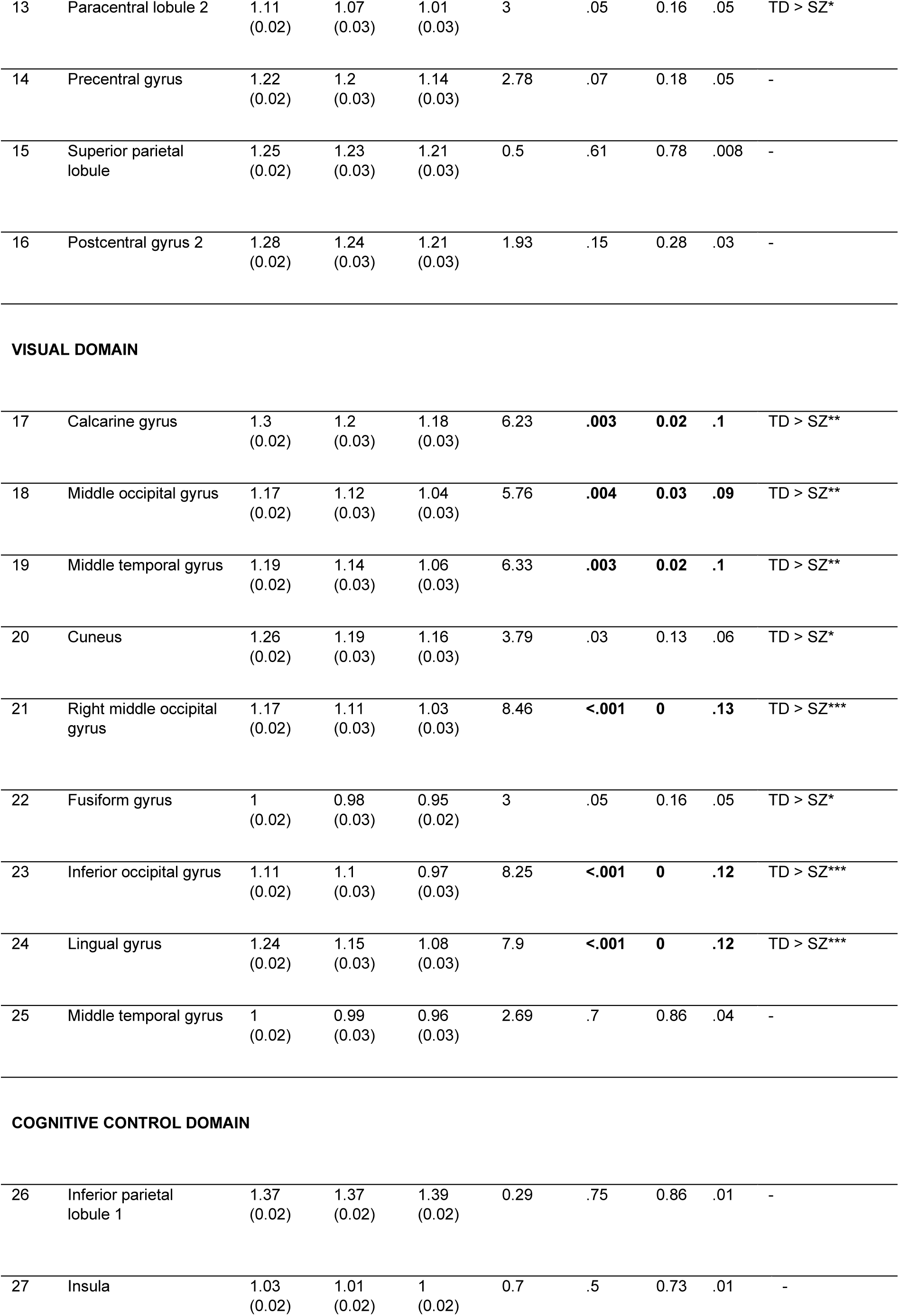
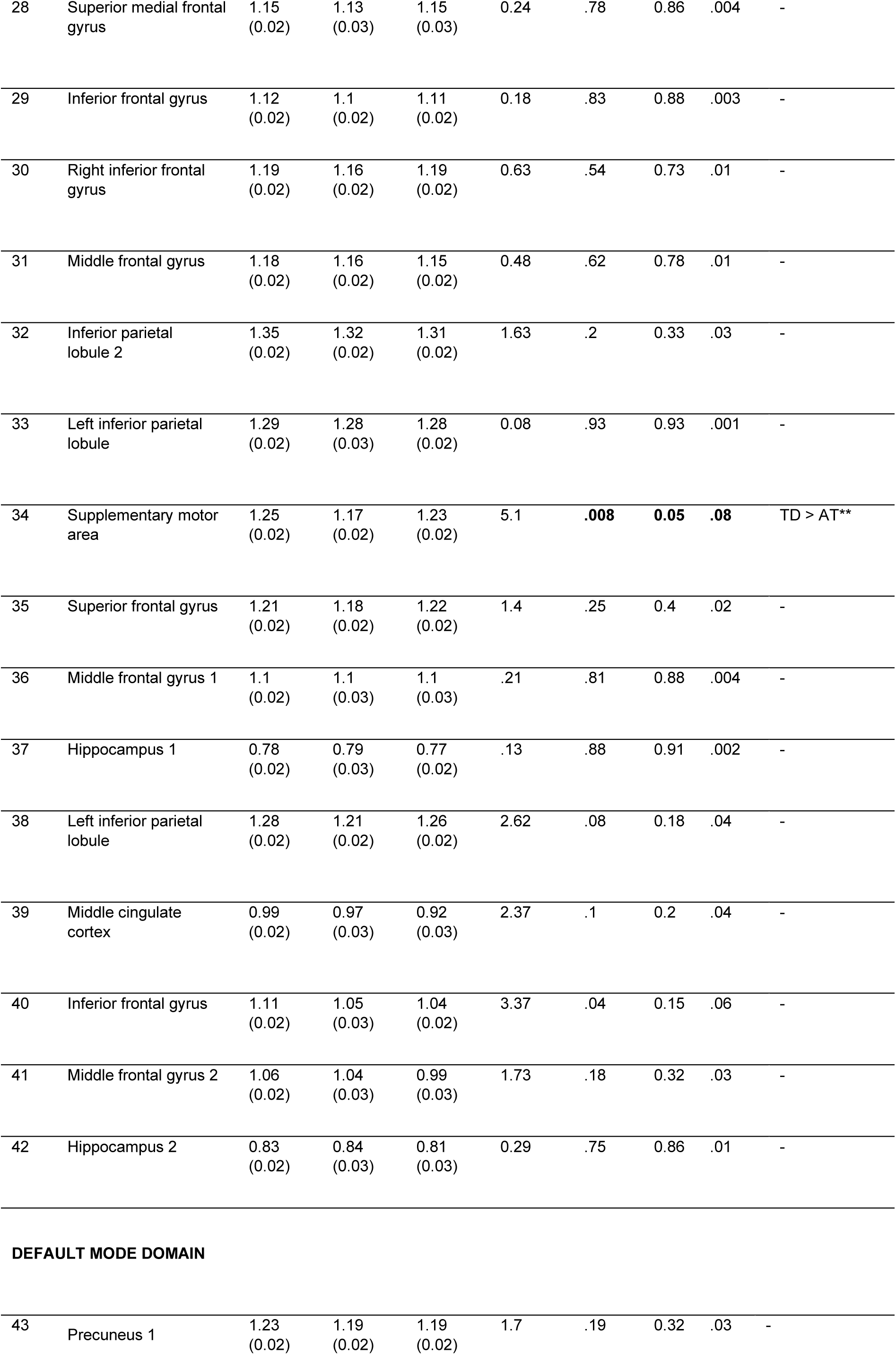
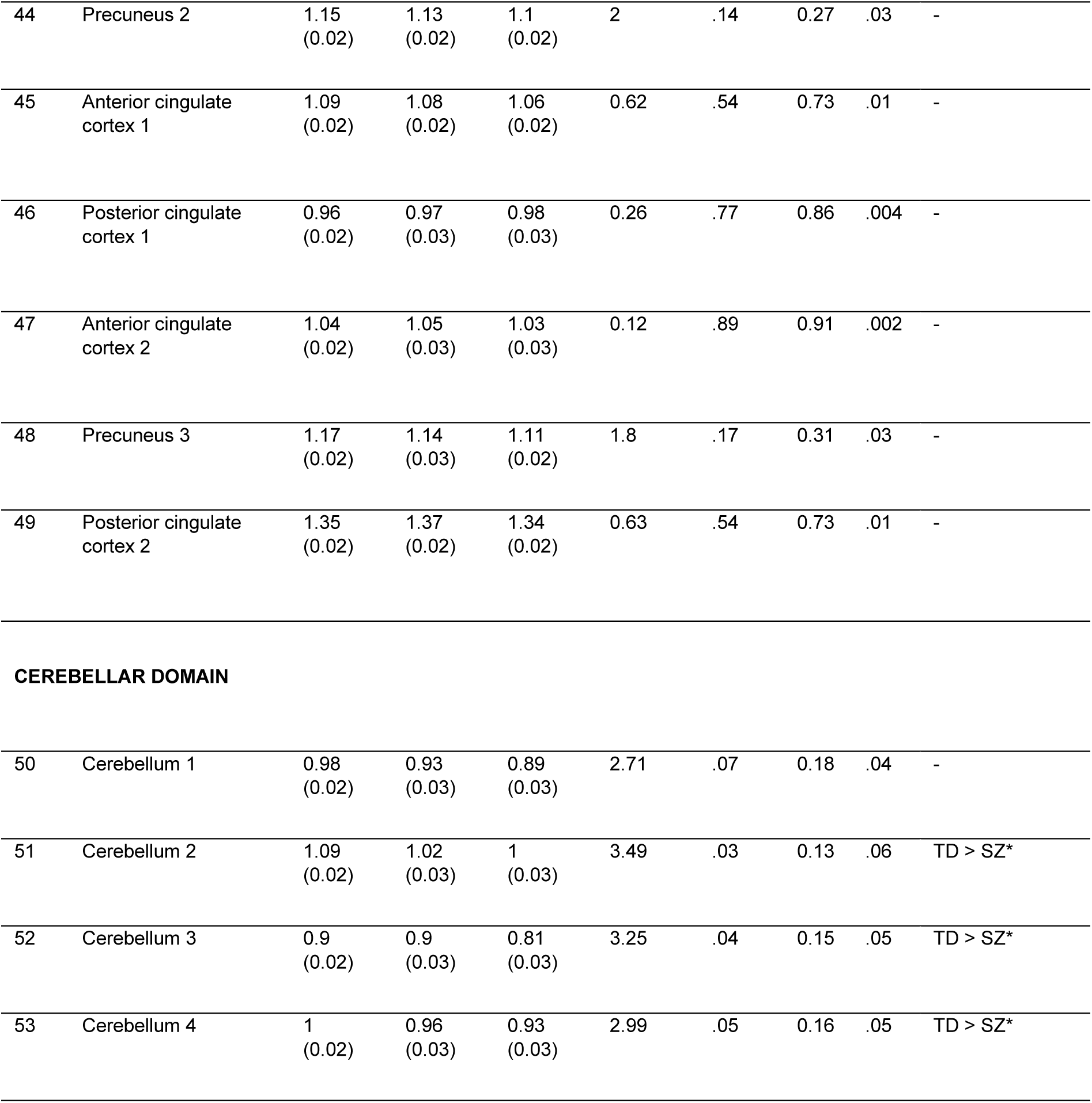
Group differences in Hurst exponent (H) per component, in the Replication dataset, computed using ANCOVA with age, sex, IQ, and FD as covariates. Multiple comparison correction of the ANCOVA p values was performed using false discovery rate (fdr). The ANCOVA effect size was calculated using partial η^2^. Post hoc tests were ran using Tukey’s HSD Test for multiple comparisons with C.I. = 95%. * *p* < .05; ** *p* < .01; *** *p* < .001

| Adjusted H mean (SE) |  |  |  |  |  |  |  |  |  |
| --- | --- | --- | --- | --- | --- | --- | --- | --- | --- |
| IC no. | | TD | AT | SZ | F<br>(2, 117) | *p* | *p<sub>tdr</sub>* | $\eta p^2$ | Post hoc |
| **SUBCORTICAL DOMAIN** |  |  |  |  |  |  |  |  |  |
| 1 | Caudate 1 | 1.1<br>(0.02) | 0.98<br>(0.03) | 0.99<br>(0.03) | 2.55 | .08 | 0.18 | .04 | - |
| 2 | Subthalamus/<br>hypothalamus | 0.8<br>(0.02) | 0.83<br>(0.03) | 0.75<br>(0.03) | 2.38 | .1 | 0.2 | .04 | - |
| 3 | Putamen | 0.98<br>(0.02) | 0.97<br>(0.03) | 0.93<br>(0.03) | 1 | .36 | 0.56 | .02 | - |
| 4 | Caudate 2 | 0.98<br>(0.02) | 0.95<br>(0.03) | 0.97<br>(0.03) | 0.01 | .6 | 0.78 | .01 | - |
| 5 | Thalamus | 0.94<br>(0.02) | 0.91<br>(0.03) | 0.85<br>(0.03) | 2.8 | .07 | 0.18 | .05 | - |
| **AUDITORY DOMAIN** |  |  |  |  |  |  |  |  |  |
| 6 | Superior temporal<br>gyrus | 0.99<br>(0.02) | 0.96<br>(0.03) | 1<br>(0.03) | 0.99 | .37 | 0.56 | .02 | - |
| 7 | Middle temporal gyrus | 0.91<br>(0.02) | 0.93<br>(0.03) | 0.93<br>(0.03) | 0.33 | .72 | 0.86 | .01 | - |
| **SENSORIMOTOR DOMAIN** |  |  |  |  |  |  |  |  |  |
| 8 | Postcentral gyrus 1 | 1.35<br>(0.02) | 1.3<br>(0.03) | 1.27<br>(0.03) | 2.55 | .08 | 0.18 | .04 | - |
| 9 | Left postcentral gyrus | 1.29<br>(0.02) | 1.21<br>(0.3) | 1.15<br>(0.03) | 6.18 | **.003** | **0.02** | **.1** | TD > SZ** |
| 10 | Paracentral lobule 1 | 1.19<br>(0.02) | 1.12<br>(0.04) | 1.04<br>(0.03) | 6.25 | **.003** | **0.02** | **.1** | TD > SZ** |
| 11 | Right postcentral<br>gyrus | 1.3<br>(0.03) | 1.25<br>(0.04) | 1.16<br>(0.03) | 4.57 | **.01** | **0.05** | **.7** | TD > SZ** |
| 12 | Superior parietal<br>lobule | 1.2<br>(0.02) | 1.16<br>(0.03) | 1.1<br>(0.03) | 2.8 | .07 | 0.18 | .05 | - |
| 13 | Paracentral lobule 2 | 1.11<br>(0.02) | 1.07<br>(0.03) | 1.01<br>(0.03) | 3 | .05 | 0.16 | .05 | TD > SZ* |
| 14 | Precentral gyrus | 1.22<br>(0.02) | 1.2<br>(0.03) | 1.14<br>(0.03) | 2.78 | .07 | 0.18 | .05 | - |
| 15 | Superior parietal lobule | 1.25<br>(0.02) | 1.23<br>(0.03) | 1.21<br>(0.03) | 0.5 | .61 | 0.78 | .008 | - |
| 16 | Postcentral gyrus 2 | 1.28<br>(0.02) | 1.24<br>(0.03) | 1.21<br>(0.03) | 1.93 | .15 | 0.28 | .03 | - |
| VISUAL DOMAIN |  |  |  |  |  |  |  |  |  |
| 17 | Calcarine gyrus | 1.3<br>(0.02) | 1.2<br>(0.03) | 1.18<br>(0.03) | 6.23 | **.003** | **0.02** | **.1** | TD > SZ** |
| 18 | Middle occipital gyrus | 1.17<br>(0.02) | 1.12<br>(0.03) | 1.04<br>(0.03) | 5.76 | **.004** | **0.03** | **.09** | TD > SZ** |
| 19 | Middle temporal gyrus | 1.19<br>(0.02) | 1.14<br>(0.03) | 1.06<br>(0.03) | 6.33 | **.003** | **0.02** | **.1** | TD > SZ** |
| 20 | Cuneus | 1.26<br>(0.02) | 1.19<br>(0.03) | 1.16<br>(0.03) | 3.79 | .03 | 0.13 | .06 | TD > SZ* |
| 21 | Right middle occipital gyrus | 1.17<br>(0.02) | 1.11<br>(0.03) | 1.03<br>(0.03) | 8.46 | **<.001** | **0** | **.13** | TD > SZ*** |
| 22 | Fusiform gyrus | 1<br>(0.02) | 0.98<br>(0.03) | 0.95<br>(0.02) | 3 | .05 | 0.16 | .05 | TD > SZ* |
| 23 | Inferior occipital gyrus | 1.11<br>(0.02) | 1.1<br>(0.03) | 0.97<br>(0.03) | 8.25 | **<.001** | **0** | **.12** | TD > SZ*** |
| 24 | Lingual gyrus | 1.24<br>(0.02) | 1.15<br>(0.03) | 1.08<br>(0.03) | 7.9 | **<.001** | **0** | **.12** | TD > SZ*** |
| 25 | Middle temporal gyrus | 1<br>(0.02) | 0.99<br>(0.03) | 0.96<br>(0.03) | 2.69 | .7 | 0.86 | .04 | - |
| COGNITIVE CONTROL DOMAIN |  |  |  |  |  |  |  |  |  |
| 26 | Inferior parietal lobule 1 | 1.37<br>(0.02) | 1.37<br>(0.02) | 1.39<br>(0.02) | 0.29 | .75 | 0.86 | .01 | - |
| 27 | Insula | 1.03<br>(0.02) | 1.01<br>(0.02) | 1<br>(0.02) | 0.7 | .5 | 0.73 | .01 | - |
| 28 | Superior medial frontal gyrus | 1.15<br>(0.02) | 1.13<br>(0.03) | 1.15<br>(0.03) | 0.24 | .78 | 0.86 | .004 | - |
| 29 | Inferior frontal gyrus | 1.12<br>(0.02) | 1.1<br>(0.02) | 1.11<br>(0.02) | 0.18 | .83 | 0.88 | .003 | - |
| 30 | Right inferior frontal gyrus | 1.19<br>(0.02) | 1.16<br>(0.02) | 1.19<br>(0.02) | 0.63 | .54 | 0.73 | .01 | - |
| 31 | Middle frontal gyrus | 1.18<br>(0.02) | 1.16<br>(0.02) | 1.15<br>(0.02) | 0.48 | .62 | 0.78 | .01 | - |
| 32 | Inferior parietal lobule 2 | 1.35<br>(0.02) | 1.32<br>(0.02) | 1.31<br>(0.02) | 1.63 | .2 | 0.33 | .03 | - |
| 33 | Left inferior parietal lobule | 1.29<br>(0.02) | 1.28<br>(0.03) | 1.28<br>(0.02) | 0.08 | .93 | 0.93 | .001 | - |
| 34 | Supplementary motor area | 1.25<br>(0.02) | 1.17<br>(0.02) | 1.23<br>(0.02) | 5.1 | **.008** | **0.05** | **.08** | TD > AT** |
| 35 | Superior frontal gyrus | 1.21<br>(0.02) | 1.18<br>(0.02) | 1.22<br>(0.02) | 1.4 | .25 | 0.4 | .02 | - |
| 36 | Middle frontal gyrus 1 | 1.1<br>(0.02) | 1.1<br>(0.03) | 1.1<br>(0.03) | .21 | .81 | 0.88 | .004 | - |
| 37 | Hippocampus 1 | 0.78<br>(0.02) | 0.79<br>(0.03) | 0.77<br>(0.02) | .13 | .88 | 0.91 | .002 | - |
| 38 | Left inferior parietal lobule | 1.28<br>(0.02) | 1.21<br>(0.02) | 1.26<br>(0.02) | 2.62 | .08 | 0.18 | .04 | - |
| 39 | Middle cingulate cortex | 0.99<br>(0.02) | 0.97<br>(0.03) | 0.92<br>(0.03) | 2.37 | .1 | 0.2 | .04 | - |
| 40 | Inferior frontal gyrus | 1.11<br>(0.02) | 1.05<br>(0.03) | 1.04<br>(0.02) | 3.37 | .04 | 0.15 | .06 | - |
| 41 | Middle frontal gyrus 2 | 1.06<br>(0.02) | 1.04<br>(0.03) | 0.99<br>(0.03) | 1.73 | .18 | 0.32 | .03 | - |
| 42 | Hippocampus 2 | 0.83<br>(0.02) | 0.84<br>(0.03) | 0.81<br>(0.03) | 0.29 | .75 | 0.86 | .01 | - |
| DEFAULT MODE DOMAIN |  |  |  |  |  |  |  |  |  |
| 43 | Precuneus 1 | 1.23<br>(0.02) | 1.19<br>(0.02) | 1.19<br>(0.02) | 1.7 | .19 | 0.32 | .03 | - |
| 44 | Precuneus 2 | 1.15<br>(0.02) | 1.13<br>(0.02) | 1.1<br>(0.02) | 2 | .14 | 0.27 | .03 | - |
| 45 | Anterior cingulate<br>cortex 1 | 1.09<br>(0.02) | 1.08<br>(0.02) | 1.06<br>(0.02) | 0.62 | .54 | 0.73 | .01 | - |
| 46 | Posterior cingulate<br>cortex 1 | 0.96<br>(0.02) | 0.97<br>(0.03) | 0.98<br>(0.03) | 0.26 | .77 | 0.86 | .004 | - |
| 47 | Anterior cingulate<br>cortex 2 | 1.04<br>(0.02) | 1.05<br>(0.03) | 1.03<br>(0.03) | 0.12 | .89 | 0.91 | .002 | - |
| 48 | Precuneus 3 | 1.17<br>(0.02) | 1.14<br>(0.03) | 1.11<br>(0.02) | 1.8 | .17 | 0.31 | .03 | - |
| 49 | Posterior cingulate<br>cortex 2 | 1.35<br>(0.02) | 1.37<br>(0.02) | 1.34<br>(0.02) | 0.63 | .54 | 0.73 | .01 | - |
| **CEREBELLAR DOMAIN** |  |  |  |  |  |  |  |  |  |
| 50 | Cerebellum 1 | 0.98<br>(0.02) | 0.93<br>(0.03) | 0.89<br>(0.03) | 2.71 | .07 | 0.18 | .04 | - |
| 51 | Cerebellum 2 | 1.09<br>(0.02) | 1.02<br>(0.03) | 1<br>(0.03) | 3.49 | .03 | 0.13 | .06 | TD > SZ* |
| 52 | Cerebellum 3 | 0.9<br>(0.02) | 0.9<br>(0.03) | 0.81<br>(0.03) | 3.25 | .04 | 0.15 | .05 | TD > SZ* |
| 53 | Cerebellum 4 | 1<br>(0.02) | 0.96<br>(0.03) | 0.93<br>(0.03) | 2.99 | .05 | 0.16 | .05 | TD > SZ* |

### 3.4. Classification accuracy and feature importance in the Exploratory dataset

Using the complete 53 Hurst feature set, the classification was: AUC = 84%, Sensitivity = 65%, and Specificity = 83% (Figure 1 A; please note that sensitivity and specificity are calculated in relation to AT).

**Figure 1.**
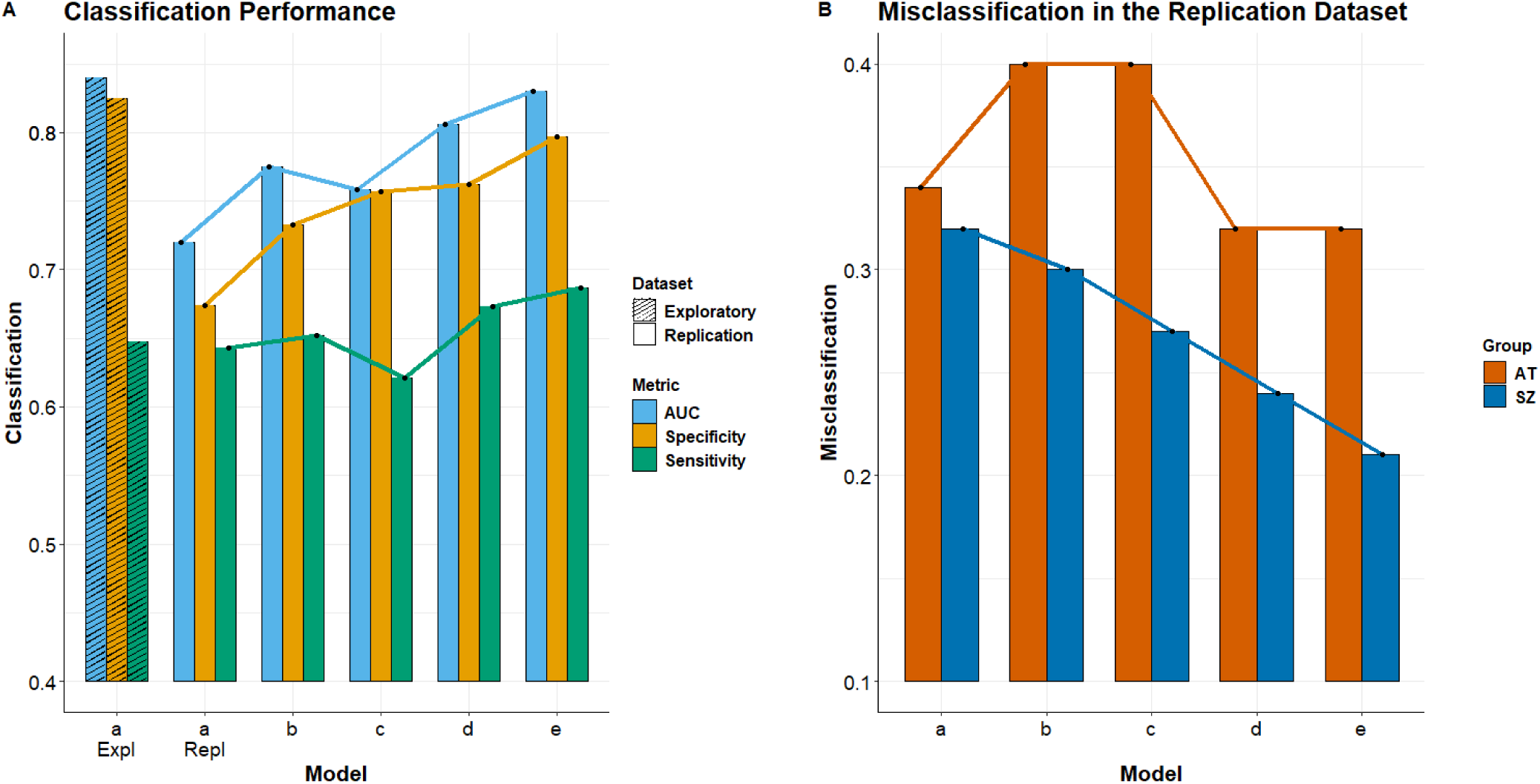
**A.** Classification performance for each model and dataset. AUC: area under the curve; a Expl: model with all 53 H values in the Exploratory dataset; a Repl: model with the 10 H values in the Replication dataset; b: model with ADOS and PANSS; c: model with ADOS, PANSS, IQ, EQ, BVAQ; d: model with the 10 H, ADOS and PANSS; e: model with the 10 H, ADOS, PANSS, IQ, EQ, BVAQ. **B.** Misclassification of participants. AT: autism; SZ: schizophrenia

Next, we used the ten ICs with the highest Hurst feature importance from the RF classification of the Exploratory dataset to simplify the feature set for RF classification of the Replication dataset. We selected these ten Hurst ICs as follows: the precuneus and the anterior cingulate cortex (i.e., ICs 43, 44, 47, and 48) from the Default Mode Network, the superior frontal gyrus (i.e., IC 35) of the Cognitive Control domain, the paracentral lobule and the precentral gyrus (ICs 10 and 14), from the Visual domain, and the middle occipital gyrus, the inferior occipital gyrus, and the lingual gyrus (i.e., ICs 18, 23, and 24), from the Sensorimotor domain (Supplement Figure 2).

### 3.5. Classification results in the Replication dataset

In the Replication dataset, we obtained the following classification performance, outlined in Figure 1A: For model (a), using the reduced Hurst feature set (10 ICs), AUC = 72%, Sensitivity = 64%, and Specificity = 67%. For model (b), using the PANSS and ADOS as features, AUC = 78%, Sensitivity = 65%, and Specificity = 73%. For model (c), using the PANSS, ADOS, EQ, IQ, and BVAQ as features, AUC = 76%, Sensitivity = 62%, and Specificity = 76%. For model (d), using the reduced Hurst feature set in combination with PANSS and ADOS, AUC = 81%, Sensitivity = 67%, and Specificity = 76%. Finally, for model (e), using the reduced Hurst feature set in combination with PANSS, ADOS, EQ, IQ, and BVAQ, AUC = 83%, Sensitivity = 68%, and Specificity = 79%.

In the Replication dataset, for each classification instance, we inspected the scaled contribution of each feature included in each of the five models. In order to account for the different number of features included in each classification model, we scaled the raw feature importance values (Supplement Figure 3) by multiplying them by the number of features included in that respective model (Supplement Figure 4). As can be seen in Supplement Figure 4, the precentral gyrus (IC 14), the inferior occipital gyrus (IC 23), the lingual gyrus (IC 24), and the precuneus (IC 48) are the most important H features overall. The most important symptom scores are the ADOS total, followed by PANSS positive and negative. However, the most important feature overall was the estimated IQ, which prompted our concern that the IQ might bias our classification output and misleadingly appear as being more important than the Hurst features in model (e), given that our groups were not matched for IQ (Table 1). We therefore repeated the classification for models (c) and (e) and found that performance remained virtually unchanged (i.e., AUC = 0.75, Sensitivity = 0.6, Specificity = 0. 76, and AUC = 0.83, Sensitivity = 0.68, Specificity = 0. 81 respectively). Following the elimination of the IQ feature from this last run of models (c) and (e), feature importance also remained unchanged compared to that shown in Supplement Figure 4.

Finally, inspection of misclassified participants (Figure 1 B), showed that on average, SZ were increasingly more accurately classified as we moved from models (a) through (e), while AT were more accurately classified when Hurst features were included (i.e., models a, d, and e).

## 4. Discussion

The current paper assessed the feasibility of using the E/I ratio, as estimated by the H exponent, to distinguish between AT and SZ. We had two main goals: (1) perform an out-of-sample replication of the classification model using a reduced set of brain-based features (i.e., the H exponent from the ten most important brain components), and (2) compare the classification accuracy when different sets of clinical, phenotypic and imaging features were combined.

We first explored group differences between the three groups, in each independent dataset (Tables 2 and 3). The most consistent findings across both datasets reflected a significantly reduced H (i.e., increased E/I) in SZ compared to TD in most areas of the cerebellar domain, the bilateral postcentral gyrus and paracentral lobule, from the sensorimotor domain, and all but one area of the visual domain. Given that levels of glutamate and GABA have been reported to vary inconsistently across SZ, and moreover, to be impacted by both medication and disease progression (see, e.g., Foss-Feig et al., 2017, for an in-depth overview), we believe that the replicability of these group differences are all the more notable. What is more, this may suggest that despite the aforementioned medication and disease progression related neurotransmitter variations in SZ, a persistently elevated E/I ratio remains, predominantly in visual processing brain areas.

Group differences were less consistent between TD and AT, and the effects mostly displayed opposite directions in the two independent datasets. Namely, in the Exploratory dataset, significantly larger H (i.e., reduced E/I) were found in AT compared to TD in the precuneus, the cerebellum, the frontal cortex, the left inferior parietal lobule, some areas of the visual domain (i.e., calcarine, fusiform, inferior occipital, and middle temporal gyrus). In the Replication dataset, the supplementary motor area showed significantly reduced H (i.e., increased E/I) in AT compared to TD. We believe that one potential explanation that could account for this reduced replicability in this case could be the increased heterogeneity of AT which may simply not have been sufficiently captured given the limited size of the Replication dataset. Indeed, Foss-Feig et al. (2017) also suggest that E/I variations appear to be more heterogenous in AT than SZ. Another aspect that could contribute to these inconsistent findings might be due to sex differences, which we were unable to assess in the current project given insufficient female AT participants. Previously, Trakoshis et al. (2020) reported that the H of the ventromedial prefrontal cortex was significantly elevated in female AT, but significantly decreased in male AT compared to TD. Thus, a more extensive exploration of sex differences in larger samples of AT could give a clearer picture on this aspect.

A direct comparison of H in AT v. SZ revealed no significant differences in the Replication dataset, but showed a consistent pattern of significantly increased H (i.e., decreased E/I) in AT compared to SZ, in all but six of the 53 brain areas. These findings however require further validation given the heterogeneity in data collection between these two groups in the Exploratory dataset.

Classification performance of AT and SZ, using the H values of all 53 brain areas, in the Exploratory dataset, was very good (AUC = 84%), with especially high specificity for SZ (83%). A similar trend was maintained when using a reduced classification model comprising the ten most important H-based features in the Replication dataset (Figure 1.A), though classification performance was overall reduced, as was to be expected in an out-of-sample replication, and given the limited size of the Replication dataset. We further explored how augmenting the feature set in the Replication dataset could improve classification performance. We found that while using PANSS and ADOS resulted in increased classification performance compared to using H alone, performance increased further when combining H, PANSS and ADOS, and was the highest when using H, PANSS, ADOS, as well as IQ, EQ and BVAQ. This steady increase in performance appeared to be mostly driven by a steady decrease in misclassified SZ (Figure 1.B). Due to some concerns that unmatched IQ between AT and SZ might be biasing these results, we re-ran these classification models without IQ as a feature, and obtained extremely similar results. We may therefore conclude that: (1) classification performance based on E/I, as estimated by H, is substantial and replicable across independent datasets, and (2) classification performance is the highest when H and clinical assessment are combined, resulting in notably decreased misclassification instances in both AT and SZ (Figure 1.B). Interestingly, AT were more frequently misclassified when only clinical assessment data were used, compared to both H only and H combined with clinical assessment (Figure 1.B). Given that both AT and SZ in the Replication dataset were acquired using the same protocol and testing environment, we surmise that inherent AT heterogeneity could best explain these different trajectories in classification performance.

While the Exploratory dataset that we used in the first step of our classification analysis offered a satisfactory amount of data, it does have the limitation that these data were collected using a variety of protocols and acquisition sites. On the other hand, while the Replication dataset offered us the possibility to test different classification models, given that identical clinical assessment protocols were used for both AT and SZ, it offered a rather small number of participants. Nevertheless, we believe that especially given these limitations, the replicated results are noteworthy.

Another limitation which we were unable to address using the currently available datasets were sex differences. This is especially true for AT, given the historic bias which has resulted in currently available samples being overwhelmingly dominated by male participants. Finally, future systematic and longitudinal studies could further clarify how H may be influenced by age (of onset), disease progression and medication in SZ.

## Data Availability

Most of the data in the current study has already been submitted to the NDAR public repository.

## Supplement

**Supplement Figure 1.**
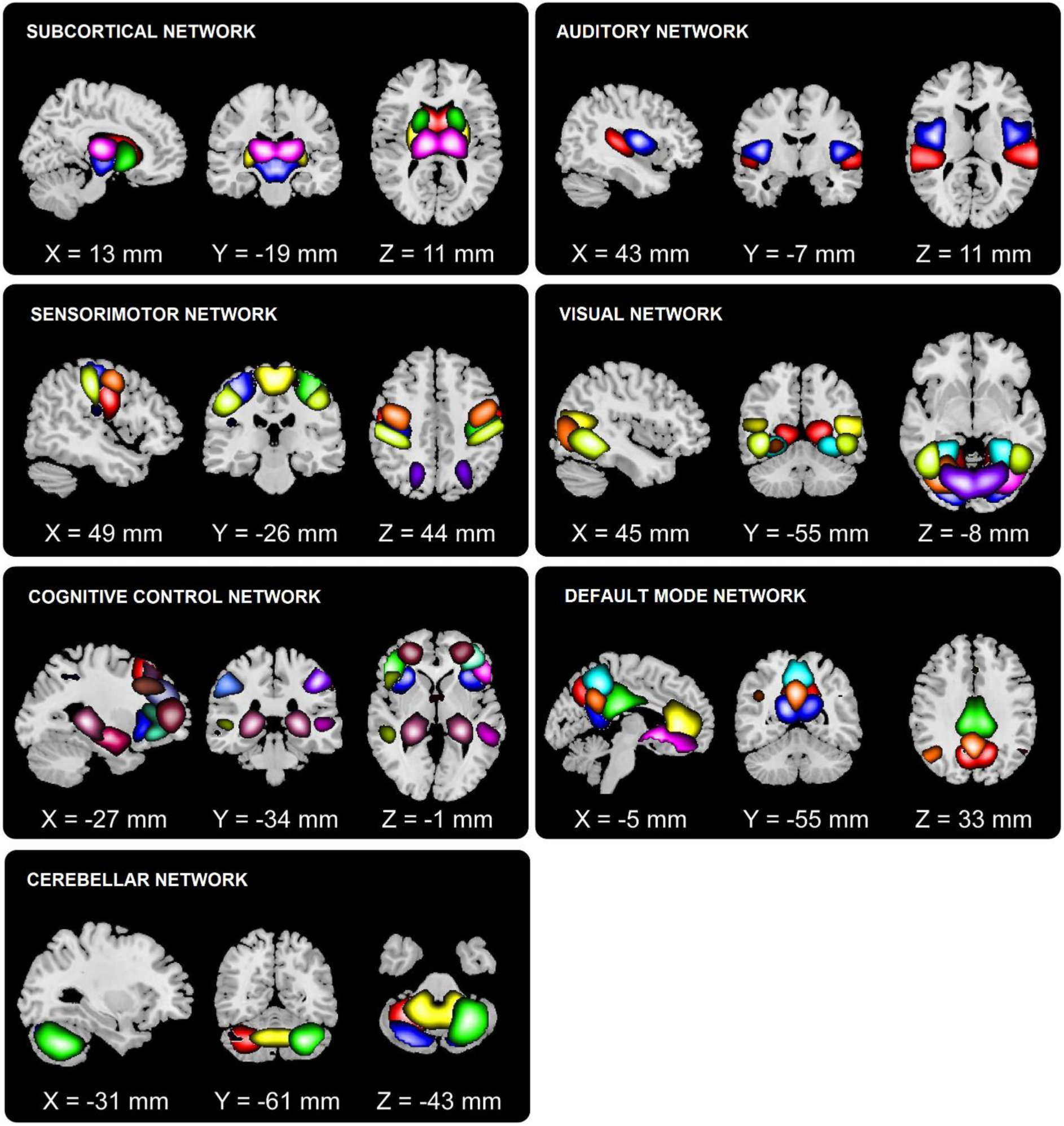
The 53 Neuromark components and corresponding domains (based on Du et al., 2020)

**Supplement Figure 2.**
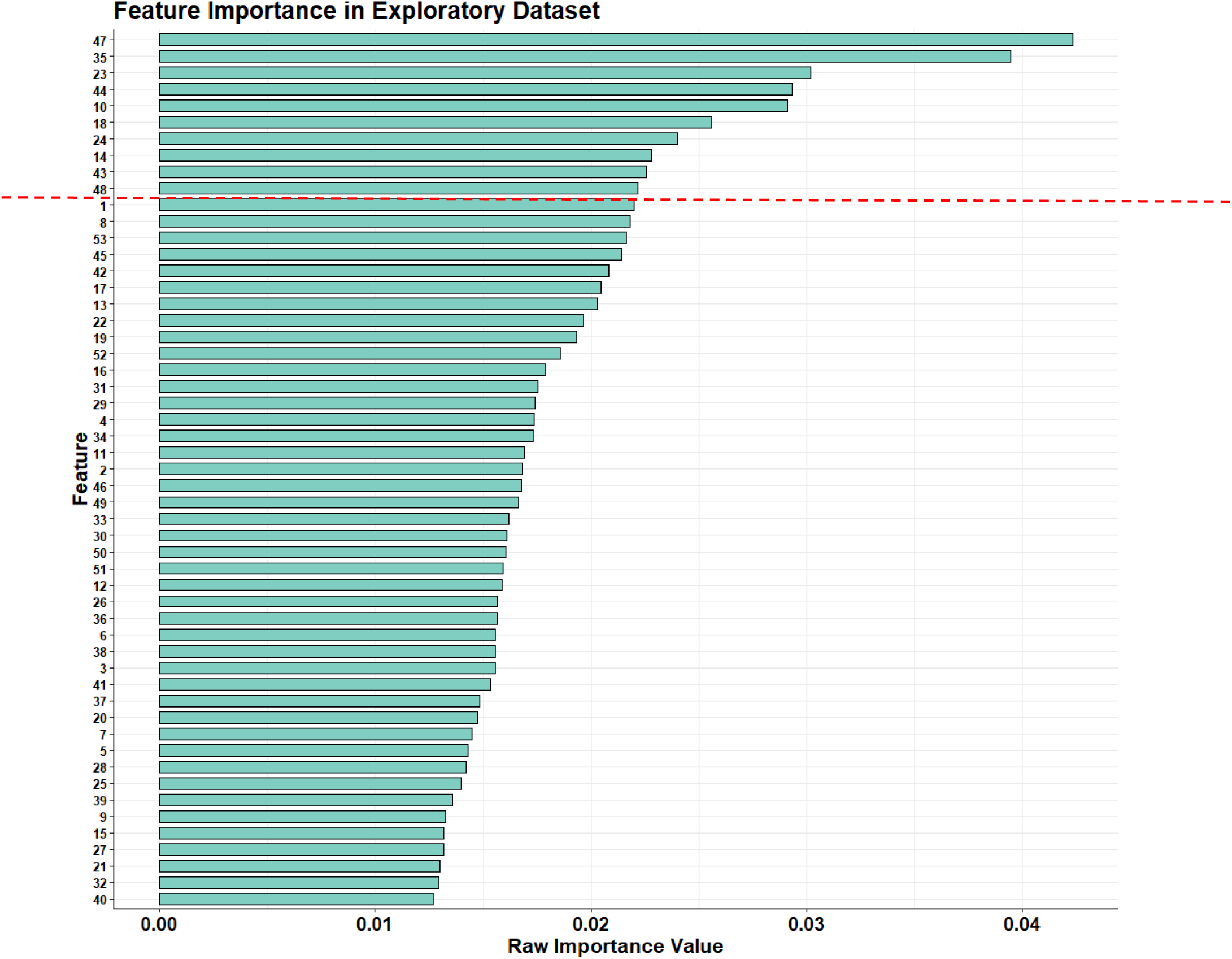
Raw feature importance values from the Exploratory dataset. The ten most important features (above the red dotted line) were selected to be used in the next classification step, using the Replication dataset.

**Supplement Figure 3.**
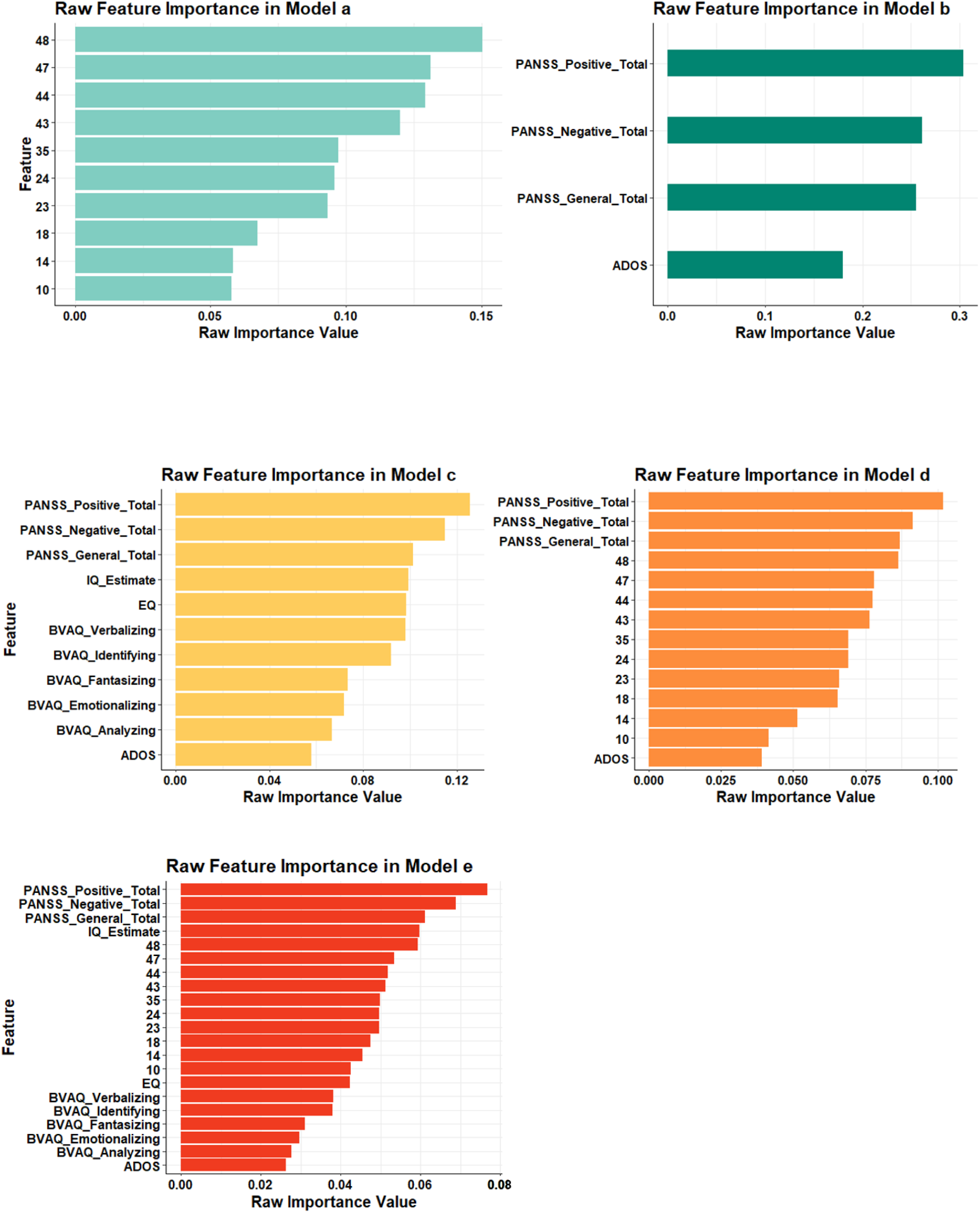
Raw feature importance values in the Replication dataset, in each of the five models. The x axis displays *raw importance values.* On the y axis, feature names are displayed; numbers correspond to the ten areas mentioned in section 3.4.

**Supplement Figure 4.**
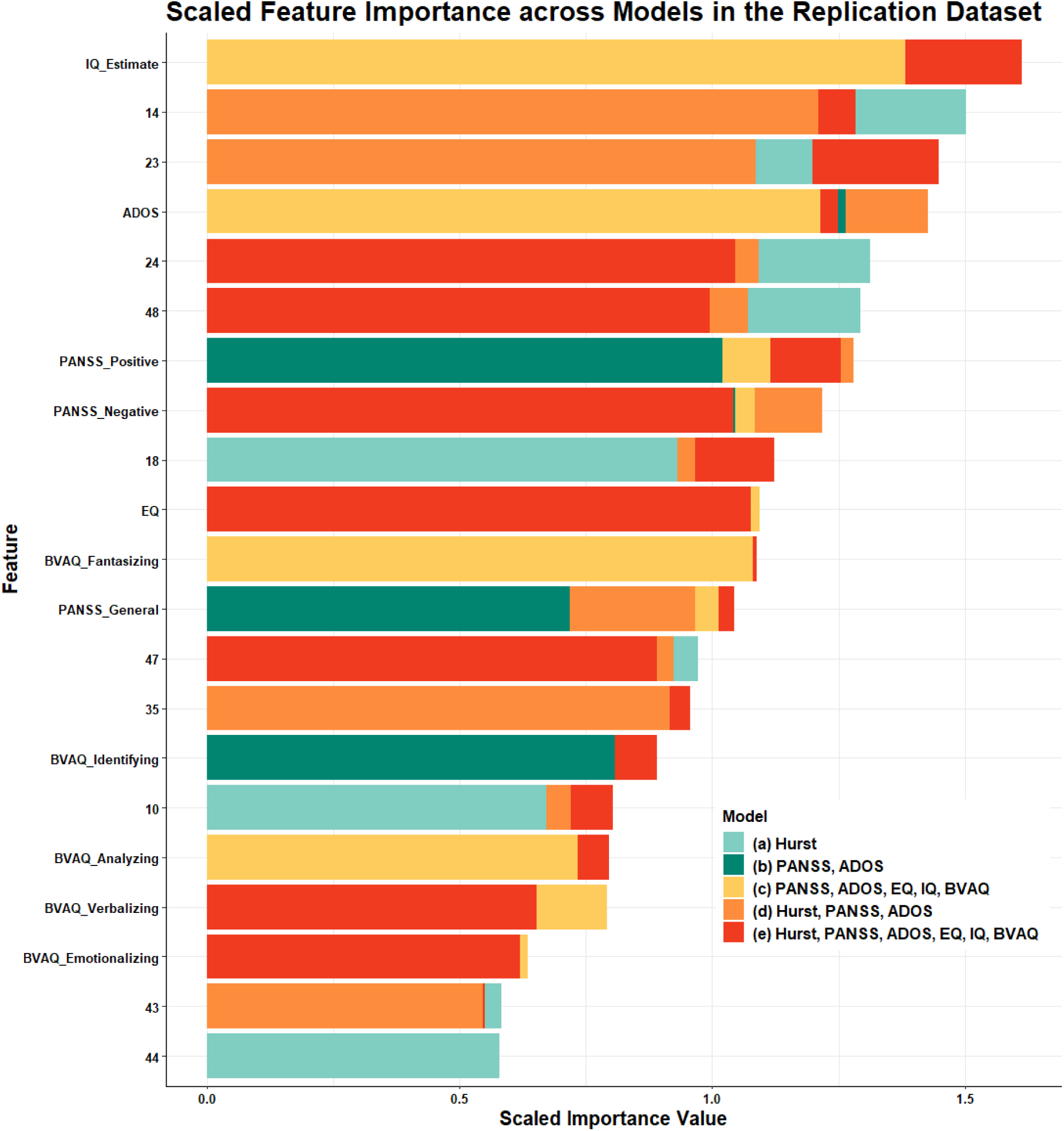
Scaled feature importance values in the Replication dataset, across all five models. The x axis displays *scaled importance values* (i.e., obtained by multiplying raw importance values by the number of features in that respective model). On the y axis, feature names are displayed; numbers correspond to the ten areas mentioned in section 3.4.

